# Profiling post-COVID syndrome across different variants of SARS-CoV-2

**DOI:** 10.1101/2022.07.28.22278159

**Authors:** Liane S. Canas, Erika Molteni, Jie Deng, Carole H. Sudre, Benjamin Murray, Eric Kerfoot, Michela Antonelli, Liyuan Chen, Khaled Rjoob, Joan Capdevila Pujol, Lorenzo Polidori, Anna May, Marc F. Österdahl, Ronan Whiston, Nathan J. Cheetham, Vicky Bowyer, Tim D. Spector, Alexander Hammers, Emma L. Duncan, Sebastien Ourselin, Claire J. Steves, Marc Modat

## Abstract

**Abstract:** *Background:* Self-reported symptom studies rapidly increased our understanding of SARS-CoV-2 during the pandemic and enabled the monitoring of long-term effects of COVID-19 outside the hospital setting. It is now evident that post-COVID syndrome presents with heterogeneous profiles, which need characterisation to enable personalised care among the most affected survivors. This study describes post-COVID profiles, and how they relate to different viral variants and vaccination status.

*Methods:* In this prospective longitudinal cohort study, we analysed data from 336,652 subjects, with regular health reports through the Covid Symptom Study (CSS) smartphone application. These subjects had reported feeling physically normal for at least 30 days before testing positive for SARS-CoV-2. 9,323 individuals subsequently developed Long-COVID, defined as symptoms lasting longer than 28 days. 1,459 had post-COVID syndrome, defined as more than 12 weeks of symptoms. Clustering analysis of the time-series data was performed to identify distinct symptom profiles for post-COVID patients, across variants of SARS-CoV-2 and vaccination status at the time of infection. Clusters were then characterised based on symptom prevalence, duration, demography, and prior conditions (comorbidities). Using an independent testing sample with additional data (n=140), we investigated the impact of post-COVID symptom clusters on the lives of affected individuals.

*Findings:* We identified distinct profiles of symptoms for post-COVID syndrome within and across variants: four endotypes were identified for infections due to the wild-type variant; seven for the alpha variant; and five for delta. Across all variants, a cardiorespiratory cluster of symptoms was identified. A second cluster related to central neurological, and a third to cases with the most severe and debilitating multi-organ symptoms. Gastrointestinal symptoms clustered in no more than two specific phenotypes per viral variant. The three main clusters were confirmed in an independent testing sample, and their functional impact was assessed.

*Interpretation:* Unsupervised analysis identified different post-COVID profiles, characterised by differing symptom combinations, durations, and functional outcomes. Phenotypes were at least partially concordant with individuals’ reported experiences. Our classification may be useful to understand distinct mechanisms of the post-COVID syndrome, as well as subgroups of individuals at risk of prolonged debilitation.

*Funding:* UK Government Department of Health and Social Care, Chronic Disease Research Foundation, The Wellcome Trust, UK Engineering and Physical Sciences Research Council, UK Research and Innovation London Medical Imaging & Artificial Intelligence Centre for Value-Based Healthcare, UK National Institute for Health Research, UK Medical Research Council, British Heart Foundation and Alzheimer’s Society, and ZOE Limited, UK.

**Research in context:** *Evidence before this study:* We conducted a search in the PubMed Central database, with keywords: (“Long-COVID*” OR “post?covid*” OR “post?COVID*” OR postCOVID* OR postCovid*) AND (cluster* OR endotype* OR phenotype* OR sub?type* OR subtype). On 15 June 2022, 161 documents were identified, of which 24 either provided descriptions of sub-types or proposed phenotypes of Long-COVID or post-COVID syndrome(s). These included 16 studies attempting manual sub-grouping of phenotypes, 6 deployments of unsupervised methods for patient clustering and automatic semantic phenotyping (unsupervised k-means=2; random forest classification=1; other=2), and two reports of uncommon presentations of Long-COVID/post-COVID syndrome. Overall, two to eight symptom profiles (clusters) were identified, with three recurring clusters. A cardiopulmonary syndrome was the predominant observation, manifesting with exertional intolerance and dyspnoea (n=10), fatigue (n=8), autonomic dysfunction, tachycardia or palpitations (n=5), lung radiological abnormalities including fibrosis (n=2), and chest pain (n=1). A second common presentation consisted in persistent general autoimmune activation and proinflammatory state (n=2), comprising multi-organ mild sequelae (n=2), gastrointestinal symptoms (n=2), dermatological symptoms (n=2), and/or fever (n=1). A third syndrome was reported, with neurological or neuropsychiatric symptoms: brain fog or dizziness (n=2), poor memory or cognition (n=2), and other mental health issues including mood disorders (n=5), headache (n=2), central sensitization (n=1), paresthesia (n=1), autonomic dysfunction (n=1), fibromyalgia (n=2), and chronic pain or myalgias (n=6). Unsupervised clustering methods identified two to six different post-COVID phenotypes, mapping to the ones described above. 14 further documents focused on possible causes and/or mechanisms of disease underlying one or more manifestations of Long-COVID or post-COVID and identifying immune response dysregulation as a potential common element. All the other documents were beyond the scope of this work. To our knowledge, there are no studies examining the symptom profile of post-COVID syndrome between different variants and vaccination status. Also, no studies reported the modelling of longitudinally collected symptoms, as time-series data, aiming at the characterisation of post-COVID syndrome.

*Added-value of this study:* Our study aimed to identify symptom profiles for post-COVID syndrome across the dominant variants in 2020 and 2021, and across vaccination status at the time of infection, using a large sample with prospectively collected longitudinal self-reports of symptoms. For individuals developing 12 weeks or more of symptoms, we identified three main symptom profiles which were consistent across variants and by vaccination status, differing only in the ratio of individuals affected by each profile and symptom duration overall.

*Implications of all the available evidence:* We demonstrate the existence of different post-COVID syndromes, which share commonalities across SARS-CoV-2 variant types in both symptoms themselves and how they evolved through the illness. We describe subgroups of patients with specific post-COVID presentations which might reflect different underlying pathophysiological mechanisms. Given the time-series component, our study is relevant for post-COVID prognostication, indicating how long certain symptoms last. These insights could aid in the development of personalised diagnosis and treatment, as well as helping policymakers plan for the delivery of care for people living with post-COVID syndrome.

## Introduction

SARS-CoV-2 is a highly infectious and rapidly mutating coronavirus. Since the first cases of human transmission in March 2020, it has infected nearly half a billion subjects worldwide^1, 2^. Early SARS-CoV-2 variants caused low respiratory tract and lung infections, with high morbidity and mortality^3^, and symptoms evolved with the emergence of new variants^4^. Many survivors report ongoing symptoms after acute illness, affecting their quality of life. Prevalence of long-term symptoms varies with viral variants^5^.

In October 2020, the definitions of ongoing COVID (OCS) and post-COVID syndrome (PCS) were introduced in the United Kingdom (UK), for otherwise unexplained signs and symptoms lasting from 4-12 weeks, or more than 12 weeks after SARS-CoV-2 infection, respectively^6^. Together OCS and PCS are often colloquially termed Long-COVID.

In June 2022, the Office of National Statistics estimated there were 2.0 million individuals reporting Long-COVID in the United Kingdom (UK)^7^, with diverse manifestations, including adverse impacts on quality of life in two-thirds^8–10^ and arising healthcare needs^11–13^. Appropriate medical and social support is thus required^14^.Previous studies have observed different presentations of PCS, and have proposed PCS should not be regarded as a unique syndrome. Several phenotypes were discerned either empirically^14–16^ or with automatic methods^17^, in relation to the causes^18–20^, phases of the disease^21^, symptom manifestation ^11, 22^, severity^9, 23, 24^, outcome^17^, and potential therapies^12^. Semi-supervised and unsupervised methods identified up to eight clusters of symptoms^17, 18, 22, 25, 26^; separating profiles of phenotypic abnormalities with enrichments in pulmonary, cardiovascular, neuropsychiatric, and constitutional symptoms such as fatigue and fever^12, 25^.

Here we hypothesised that, among the heterogeneous presentation of PCS, distinct phenotypes and symptom profiles might be identified, potentially due to differing pathophysiology. Moreover, we hypothesised that PCS heterogeneity may vary with SARS-CoV-2 variants and an individual’s vaccination status at the time of infection. Lastly, we hypothesised that identifying discrete illness profiles conveys clinical usefulness that could guide treatment approaches. Our work thus aims to inform personalised management and prognostication of PCS.

To these ends, we conducted an unsupervised clustering analysis of symptoms from community-based individuals with PCS, drawn from a uniquely large population with prospective self-reported data, collected over the course of the pandemic in the UK. Clustering was performed independently for three viral variants, and vaccination status at the time of infection; and clusters were verified against individual self-assessment of the impact of PCS in their daily routines.

## Methods

### Data

The data in this study were prospectively collected in the COVID Symptoms Study (CSS), using a mobile health application developed by ZOE Limited, in collaboration with scientists from King’s College London, Massachusetts General Hospital, Uppsala and Lund University, Sweden ^27, 28^.

Briefly, after providing informed consent, participants self-reported symptoms (both free text and in response to targeted questions) (Supplementary Table 1), SARS-CoV-2 testing and results, vaccination status, health care access, and demographic and co-morbidity data. The initial list of 20 explicit questions regarding symptoms expanded after 4 November 2020 to 34 symptoms, in response to evolving knowledge of COVID-19 infection (Supplementary Table 1). Although participants can also proxy-report for others, the current study was limited to self-reported information.

We defined individuals with PCS as those reporting symptoms for longer than 84 days (12 weeks), as per the UK National Institute for Health and Care Excellence definition^6^. We defined Long-COVID as all individuals reporting symptoms for longer than 28 days (four weeks). As this study focused on PCS, we did not perform formal modelling regarding the profile of Long-COVID individuals, merely descriptive analyses of the demographic profile of those individuals.

To study the effect of vaccination on PCS symptom clustering, we split the data into vaccinated and unvaccinated individuals at the time of their SARS-CoV-2 infection. Post-vaccination infection was defined as a positive test at least seven days after first vaccination dose. Participants reporting a positive test within seven days of a dose of vaccination were excluded, as they are unlikely to have protective effects of vaccination within this timeframe^29–31^. Time since vaccination (beyond seven days) and vaccination dose number were not considered in the analyses.

Data were collected and analysed separately for three periods of the pandemic in the UK, defined according to the predominant circulating SARS-CoV-2 strain (>=80% of tested samples caused by the same strain), documented by the COG-UK/Mutation Explorer ^32^ (Supplementary Figure 1). The period of wild-type predominance in the UK lasted from 24^th^ March 2020 to 29^th^ November 2020, alpha from 10^th^ January 2021 to 25^th^ April 2021, and delta from 26^th^ May 2021 to 8^th^ December 2021.

Using ExeTera software^33^, data were extracted and curated for UK individuals meeting the following inclusion criteria: (1) a positive SARS-CoV-2 test (either reverse transcription-polymerase chain reaction (PCR) or lateral flow antigen test (LFAT)), between 24^th^ March 2020 and 8^th^ December 2021 and within one of the windows with a predominant circulating SARS-CoV-2 strain; (2) with at least four weeks of logging as healthy (i.e. reporting “I feel physically normal”) preceding a test, excepting the immediate five days prior to test, (3) illness duration (i.e. symptoms reported) for at least 28 days post-diagnosis (we considered up to five days before a positive test outcome to include the symptoms at the onset of the illness), (4) symptom logging at least once weekly until reporting feeling healthy for more than four weeks, as previously^34^. Exclusion criteria included: (1) subjects with inconsistent non-physiological information (e.g., body mass index (BMI) values below 10 or above 95 kg/m^2^); (2) proxy-reported individuals; (3) age below 18 or above 100 years old; (4) participants reporting a positive test within seven days of a dose of vaccination. Figure 1 details the study design and its inclusion and exclusion criteria.

**Figure 1.**
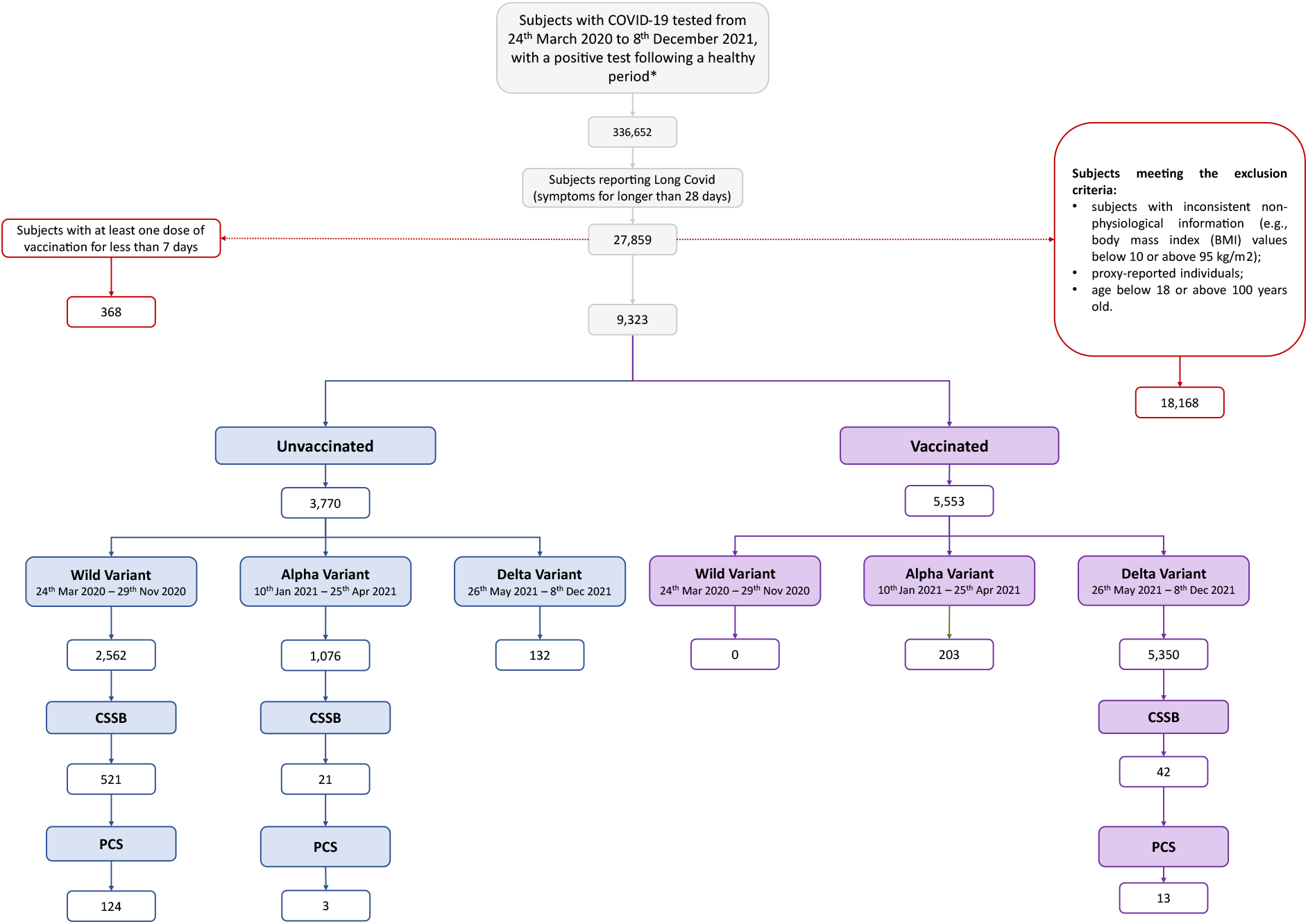
Flowchart of participants satisfying the inclusion and exclusion criteria for this study. The candidates were included the study if reporting a positive test following logging as healthy for at least 4 weeks (*). Individuals who reported symptom duration for at least 28 days were then retained according to inclusion/exclusion criteria. Individuals were further subdivided according to the timing of SARS-CoV-2 infection relative to vaccination: vaccinated (considered from at least one week after the first dose) and unvaccinated (infection prior to the first dose of vaccine). Parsing of individuals by variant type is defined by positive testing during time periods where the prevailing UK strain was >=80 % of circulating SARS-CoV-2, using UK-COG data (see text for details).CSSB: COVID Symptoms Study Biobank; PCS: post-COVID syndrome.

In addition to data logged through the ZOE COVID Study App, further data were available on a subset of participants who had been recruited to a biobank (the COVID Symptom Study Biobank [CSSB] ^35^) between October 2020 and April 2021. Participants in the CSSB responded to an online questionnaire interrogating the impact of ongoing symptoms on daily life (considered retrospectively), as part of investigations regarding the effects of the pandemic on mental and physical health. Data responses from a subset of these questions (Supplementary Table 2) were included in the present study, to assess for an association between any identified clusters and the daily living of PCS individuals.

The CSS data were split into training and testing sets for, respectively, (1) symptom cluster optimisation; and (2) evaluation of the clinical relevance of any observed clusters and cluster reproducibility in a new population. The testing set included 50% of the CSSB population and a 1:1 matched population from the CSS app who were not recruited to the Biobank. The training set comprised the remaining 50% CSSB population and all remaining eligible CSS app participants (sample size description available in Figure 1).

### Statistical analysis

We performed a Mann-Whitney test (alpha = 0.05, before multiple corrections) to assess whether differences in symptom prevalence existed between vaccinated and unvaccinated populations for alpha and delta variants, independently. Wild-type was not formally accessed due to the lack of data of vaccinated individuals infected with this variant.

### Data analysis and clustering estimation

Prior to clustering analysis, three clinicians independently categorised symptoms, into symptom domains, for descriptive purposes. Discrepancies in allocation of domains were then discussed and resolved jointly. The symptom domains were upper respiratory, abdominal, cardiorespiratory, immune-related/cutaneous, central neurological, and systemic/inflammatory. This grouping was then used to inform visualise and describe symptom cluster profiles; but was not used otherwise in the model.

Using an unsupervised time series clustering approach, we characterised symptom profile(s) for individuals with PCS, for vaccinated and unvaccinated populations, and for each variant of the virus. We estimated the symptom time series per patient using symptom aggregation per week – i.e., we computed the frequency of each symptom per week as the sum of its reports during seven days. We treated the missing reports within a week as missing data; thus, missing symptom reports were linearly interpolated independently per symptom within each week. We adopted a model, previously used on individuals with COVID-19^34^, which relies on the Multivariate time-series Clustering based on Principle Component Analysis (Mc2PCA) method^36^ to estimate symptom clusters. This method allows the clustering of time series with distinct duration by establishing the comparisons between the samples using covariance matrices, thus enabling the comparison of subjects with heterogeneous symptom reporting. The number of clusters was optimised through an iterative process using a K-means approach. For each cluster, the projection was computed using the singular value decomposition (SVD) of the average of the covariance matrices, where the first 6 dimensions were used. The process was repeated until the convergence criterion was reached: the error after projection is below 10^-4^. The optimal number of clusters, hereafter defined as optimised clusters, per variant and per population (vaccinated *versus* unvaccinated) was found through the Bayesian Information Criterion (BIC), considering the global minimum, or the first local minimum, alternatively, obtained across 10 random initialisations. A bootstrapping analysis was conducted for the wild-type variant (50 randomly selected samples for training and testing respectively) to assess the robustness of the number of optimised clusters. For alpha and delta, only 10 bootstraps were performed given the reduced sample size of their testing set (limited number of CSSB samples).

Given the small numbers of unvaccinated individuals in the alpha and delta periods, we were not able to perform clustering of symptoms for these populations.

### Clustering assessment and phenotype profiling

The best performing clusters across bootstrapping (i.e., the samples with the lowest BIC and error), were used to assess the profile of the different clusters (Supplementary Figure 2).

We computed the profile of the optimised clusters, per variant, as the average interpolated frequency of the symptoms during illness. Additionally, we assessed the proportion of subjects presenting with each symptom per week. To differentiate the symptom profile per cluster, we conducted a significance analysis: we computed the z-score between the mean symptom profile across all subjects, independently of the cluster, and the average profile of subjects belonging to a given cluster. The symptoms above the 3^rd^ quartile of the median z-scores during the illness were identified as the most relevant within each cluster. As a result, we characterised each cluster using the set of symptoms identified by the significance analysis as a proxy for the distinctive illness phenotype within each cluster.

To assess if the average profile per cluster was affected by intra-cluster variability, we compared the average profiles to an example subject, selected as the individual with the minimum distance to the centroid of the cluster, among a sub-population with age and BMI within mean ± one standard deviation of the cluster population.

Lastly, we assessed differences in demographic profiles of subjects (age, BMI, and number of comorbidities (Supplementary Table 1)) across clusters for the same variant and population, using the Mann-Whitney test.

### Long-lasting effects and clinical validation

To assess the clusters’ generalisation to different populations, we computed the cluster prediction for an independent sample of individuals infected during the wild-type period who had additional data (CSSB Long-COVID questionnaire retrospective data), retrospectively collected between May and June 2021 (Supplementary Table 2). Additionally, to assess the clinical relevance of optimised clusters and any impact on daily living, we included information on the impact of symptoms on daily life, health care usage (both acutely and >12 weeks from infection) and re-infection prior to May-June 2021.

We applied a multivariate generalised linear regression model (GLM), here a logistic regression, to study the effect of each cluster on participants’ outcomes, assessed through the follow-up questionnaire. We considered age, gender, BMI, and cluster classification as covariates of the model. Odds ratios were calculated between the predictor, which consisted of questionnaire answers binarised as reported impact (1) and no impact (0), and the clusters (one-hot-encoded). Note that different models were optimised for the different questions.

### Ethical approval

The app and CSS were approved in the UK by KCL’s ethics committee (REMAS no. 18210, review reference LRS-19/20–18210). All app users provided informed consent for use of their data for COVID-19 research. The CSSB was approved by the NHS Health Research Authority (reference 20/YH/0298) and licensed under the Human Tissue Authority (reference 12522). CSSB participants were invited to join from the app user base and provided informed consent to participate in the additional questionnaire and sample collection studies, and for linkage to app-collected data. Furthermore, this project was discussed with individuals with lived experience of PCS and their families through the CSSB volunteer advisory panel at the beginning of the project and during the analysis phase. All research was conducted in full compliance with the Declaration of Helsinki and its updates.

## Results

### Cohort description

The dataset comprised a total of 9,323 subjects overall from CSS, (Figure 1). Among the overall CSS population, 584 also belonged to the CSSB, of which 140 reported PCS (Figure 1, Table 1).

**Table 1.**
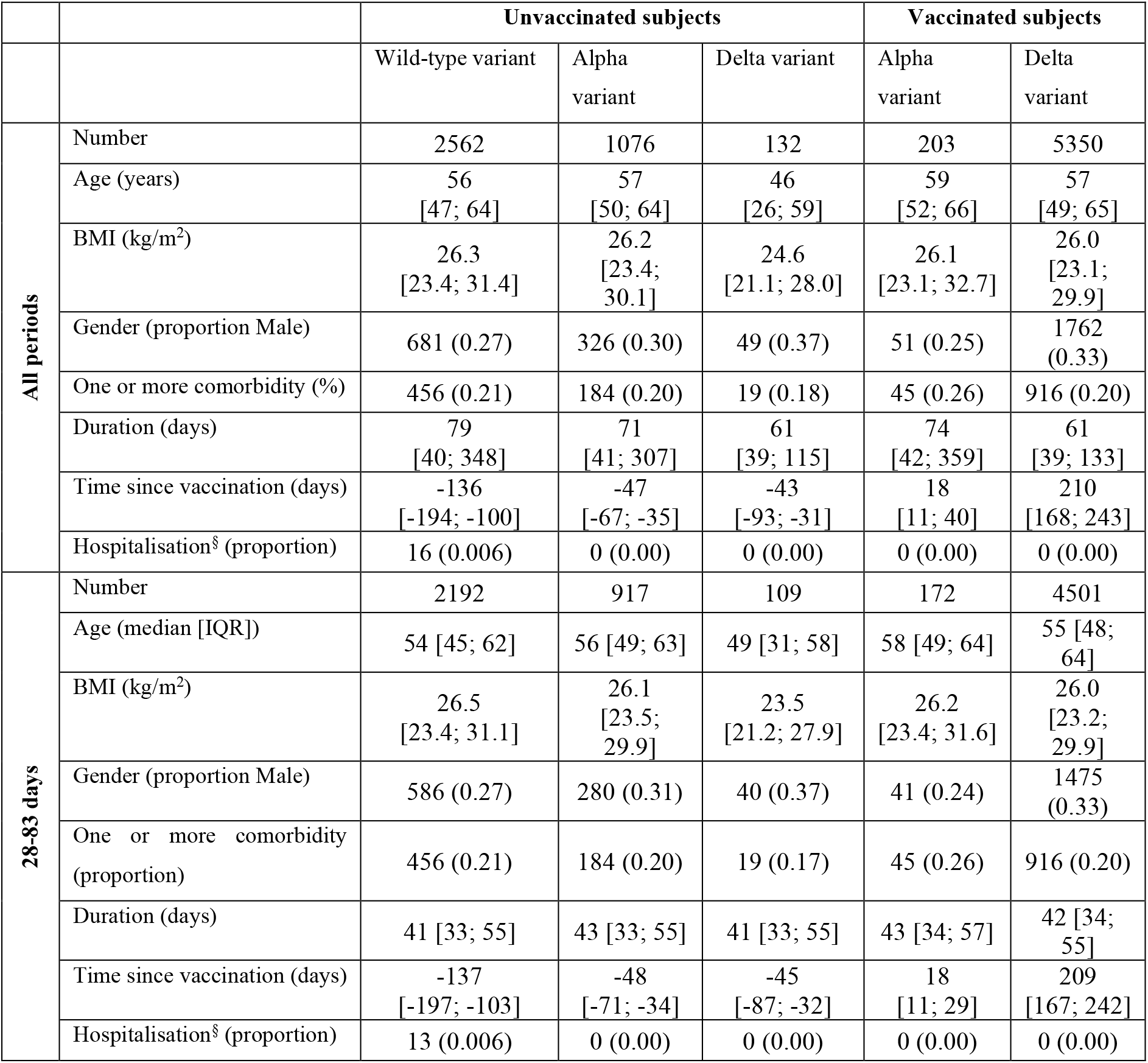
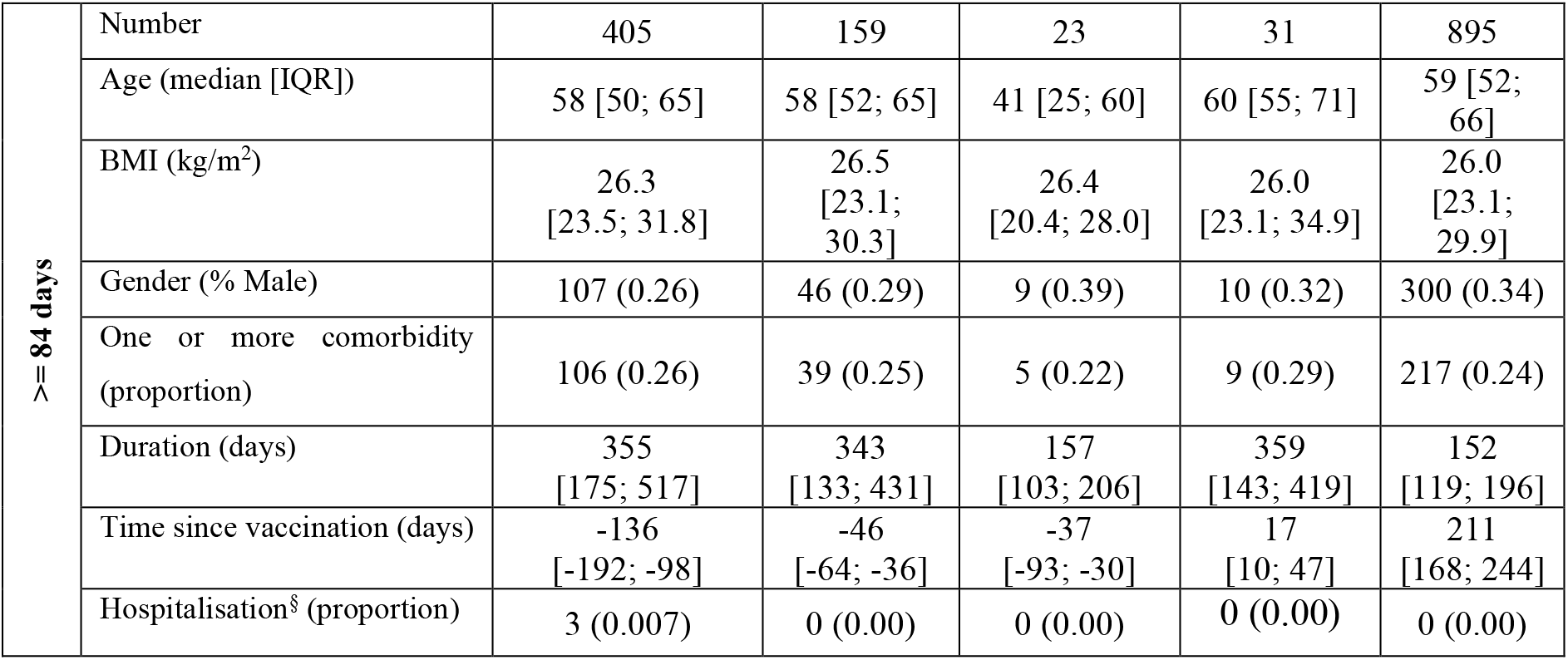
Demographic information of the study population. Median values and interquartile range [IQR] are presented for age, body mass index (BMI), duration, and time since vaccination. Gender, individuals with more than one comorbidity, and hospitalisation (§ during this period of illness) are presented as proportions.

We assessed differences in symptom prevalence and duration for individuals reporting PCS before and after vaccination (Figure 2, Supplementary Table 3). Vaccination was not available to the general community during the wild-type variant period; consequently, wild-type was not considered in this particular analysis. There were no significant differences across populations, suggesting that symptom prevalence among individuals developing PCS does not depend on the timing of infection relative to vaccination status (Supplementary Table 3).

**Figure 2.**
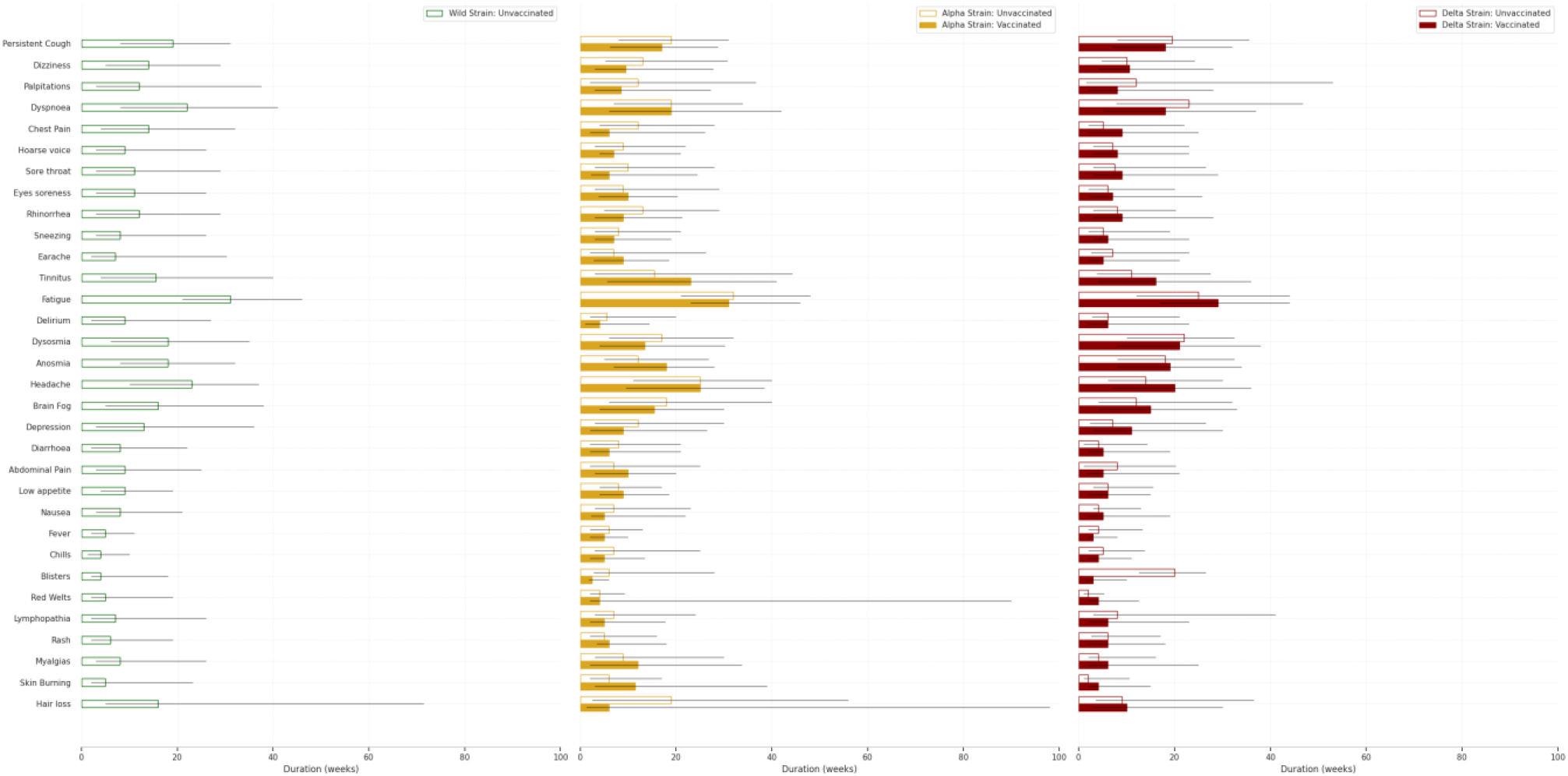
Symptom prevalence and duration for subjects reporting PCS. Median duration of symptoms, in weeks, for subjects reporting infection before vaccination (unvaccinated – hollowed bars; subsequently vaccinated – filled bars; interquartile range (IQR) represented by the black lines; green: wild-type variant; yellow: alpha variant; red: delta variant).

### Clustering assessment and phenotype profiling

We assessed the symptom profile per variant and per population (vaccinated *vs* unvaccinated) in individuals with PCS illness. For individuals infected with wild-type variant, we only considered twenty symptoms (reflecting the twenty direct questions in the CSS app during this period); for both alpha and delta variants, we considered all thirty-four symptoms available (Supplementary Table 1).

Overall, we found differing numbers of clusters per variant (Figures 3, 5 and 7), each with specific illness duration and symptom profile (both prevalence and duration). The optimised number of clusters was determined via BIC minimisation (Supplementary Figure 2). A detailed description of cluster sizes and their symptom profiles and duration, per variant, is presented in Supplementary Table 4. These results suggest that PCS may be heterogeneous, depending on SARS-CoV-2 variant.

Nonetheless, based on symptom relevance (highest ranked z-score symptoms – Figures 4, 6 and 8, Supplementary Figures 3, 6 and 9), we also found three common patterns across variants: (1) a central neurological cluster, often without a great impact of other symptoms (alpha and delta variants), (2) a cardiorespiratory cluster, typically linked to severe symptoms, such as severe dyspnoea (nb.: only mild to moderate dyspnoea for the wild-type variant), and (3) a systemic/inflammatory cluster also often manifesting other immune-related symptoms. Additionally, abdominal symptoms were often predominantly isolated within a single (smallest) cluster, evident across the three variants. Lastly, the immune-related and cutaneous symptoms predominantly manifest within a single cluster, with some minor occurrences of these symptoms within other clusters. When present, immune-related symptoms showed a high relevance, contributing for cluster uniqueness, but a low prevalence among the individuals, and often co-occurred with systemic and inflammatory symptoms.

#### PCS clusters from infection with wild-type variant

We identified four symptom clusters for subjects infected by the wild-type variant (Figure 3). The largest cluster, cluster wild-A (n=138, 51%) was characterised by upper respiratory and central neurological symptoms. The next largest, cluster wild-B (n=60, 22%), was heterogeneous and included symptoms relating to immune, central neurological, and respiratory, systemic/inflammatory systems. This cluster had the most severe symptoms including prolonged fever, and 20% of individuals had dyspnoea affecting their ability to function normally eight weeks post-infection. Cluster wild-C (n=38, 14%) contained predominantly upper respiratory symptoms, particularly hoarse voice, as well as anosmia and low appetite. Lastly, cluster wild-D (n=37, 14%) exhibited abdominal symptoms as well as central neurological (headache and anosmia), and upper respiratory (sore throat) symptoms also common to other clusters. Together with cluster wild-B, this cluster had a longer duration and greater symptom severity. Age, gender, and BMI were not different between wild-type clusters (Supplementary Figure 4). Supplementary Figure 5 presents the individual symptom profile for the optimised clusters.

**Figure 3.**
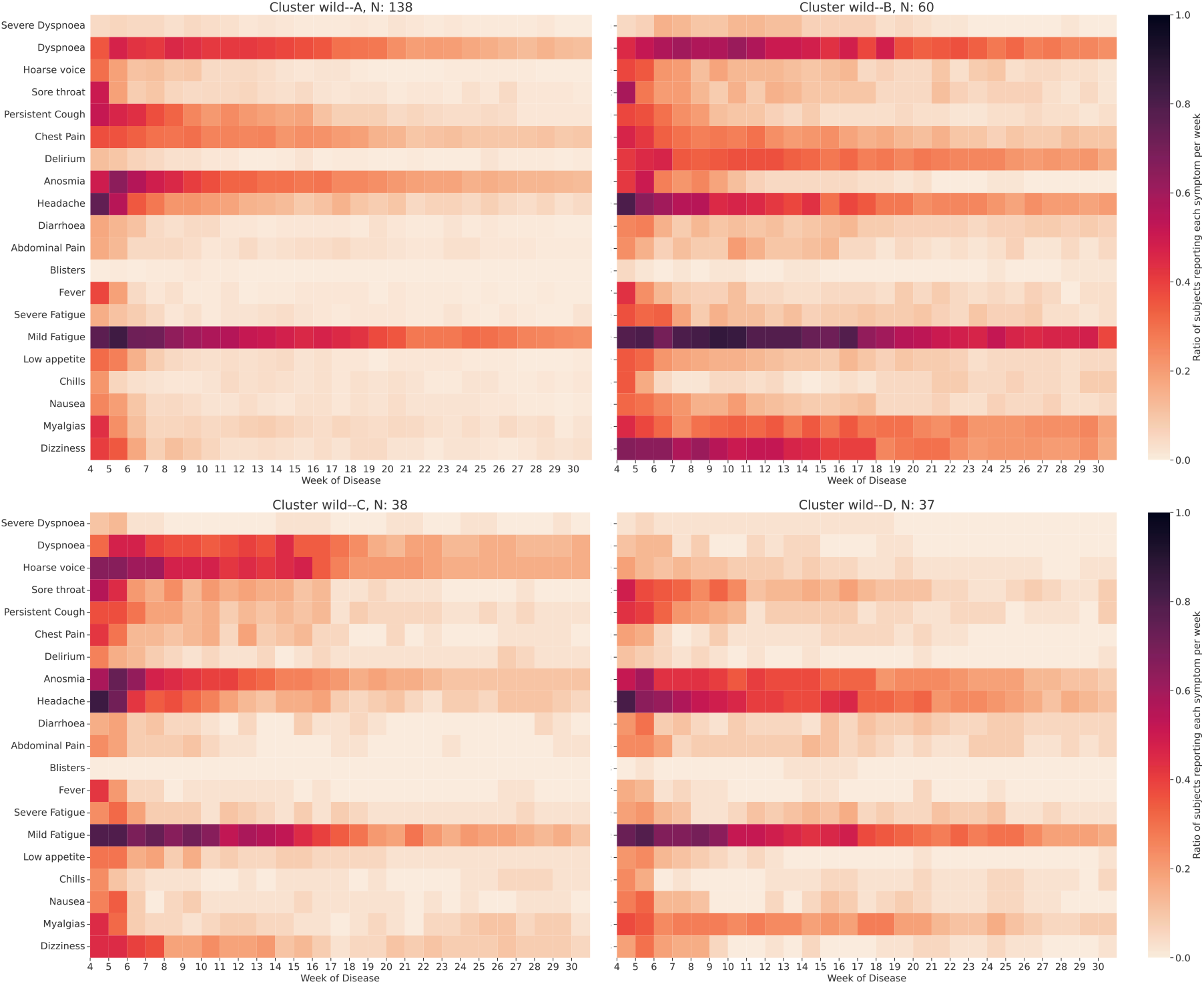
Symptom prevalence profile for PCS clusters for the wild-type variant, for subjects with a symptomatic illness longer than 84 days. The proportion of subjects reporting each symptom (ratio) per week is encoded by the colourmap (darker represents a higher ratio).

**Figure 4.**
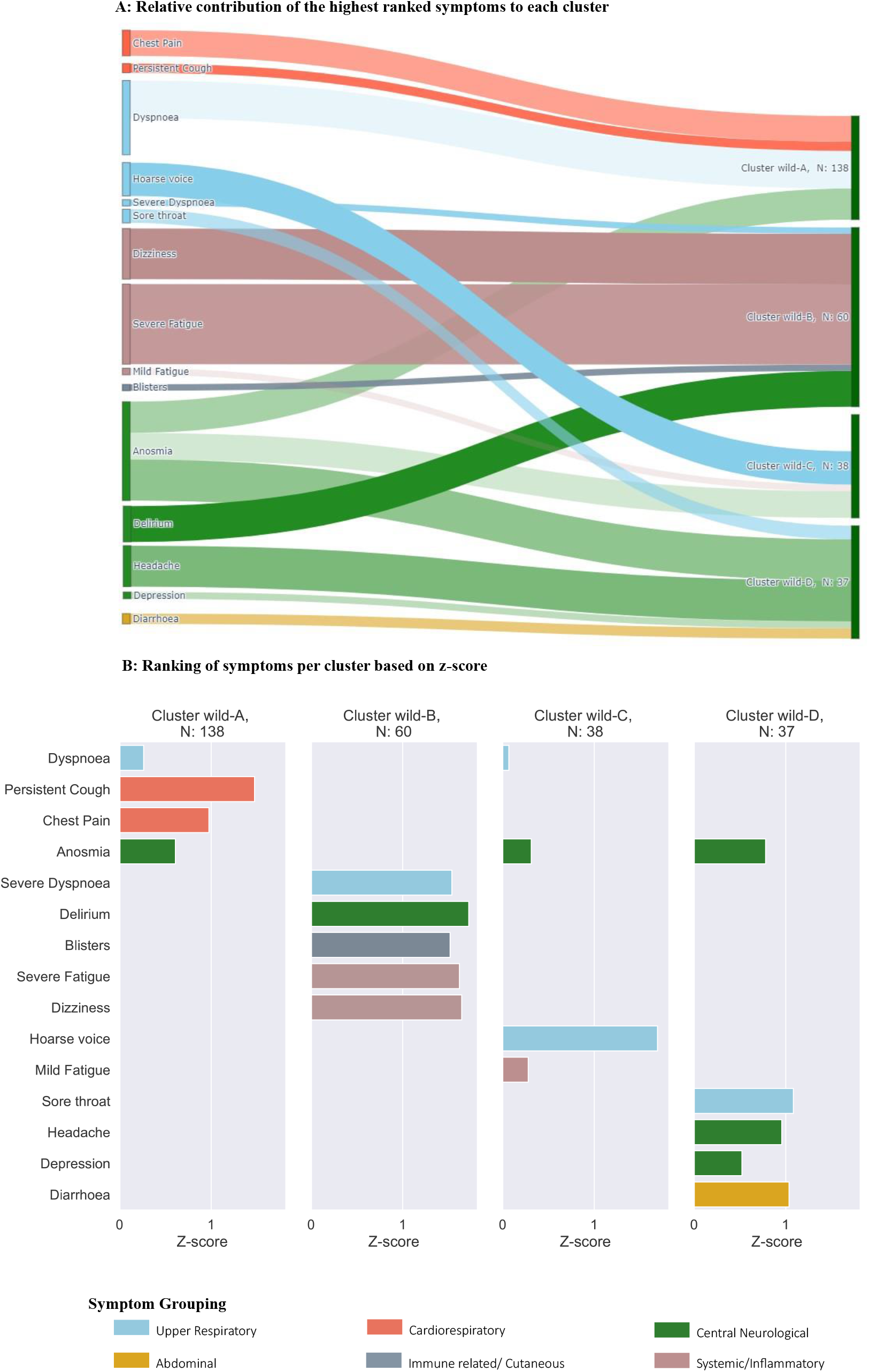
Relative contribution of symptoms per cluster for PCS in individuals with the wild-type variant. Symptoms are selected based on the 3^rd^ quartile of the z-scores (only positive z-scores are shown), aiming to highlight the contributing most symptoms in each cluster. **A:** relative contributions of symptoms per cluster. The width of shadow connections encodes the symptom prevalence (median number of individuals reporting the symptom within the cluster), while colour intensity encodes the relevance of symptoms (z-score) per cluster. **B:** Ranking of symptoms based on z-scores, per cluster.

#### PCS Clusters from infection with the alpha variant

We identified seven clusters among unvaccinated PCS subjects infected with the alpha variant (Figure 5). The largest cluster, cluster alpha-A (n=47, 30%), was dominated by anosmia, whereas the smallest, cluster alpha-G (n=13, 8%), was highly heterogeneous and polysymptomatic (Figure 6, Supplementary Figure 6). The individualised patterns reflected again consistency, matching the average symptom profiles within each alpha cluster (Supplementary Figure 7). Similar to the results for the wild-type variant, abdominal symptoms were mainly present within a single cluster, cluster alpha-F. The clusters differed in median illness duration (Supplementary Table 4), with the shortest duration for cluster alpha-B (16.5 [14; 28.75] weeks, predominantly upper respiratory symptoms) and the longest in cluster alpha-E (23 [13.75; 41] weeks, predominantly central neurological symptoms). We did not find differences in demographic features (age, BMI) within or across clusters, after correction for multiple comparisons (Supplementary Figure 8).

**Figure 5.**
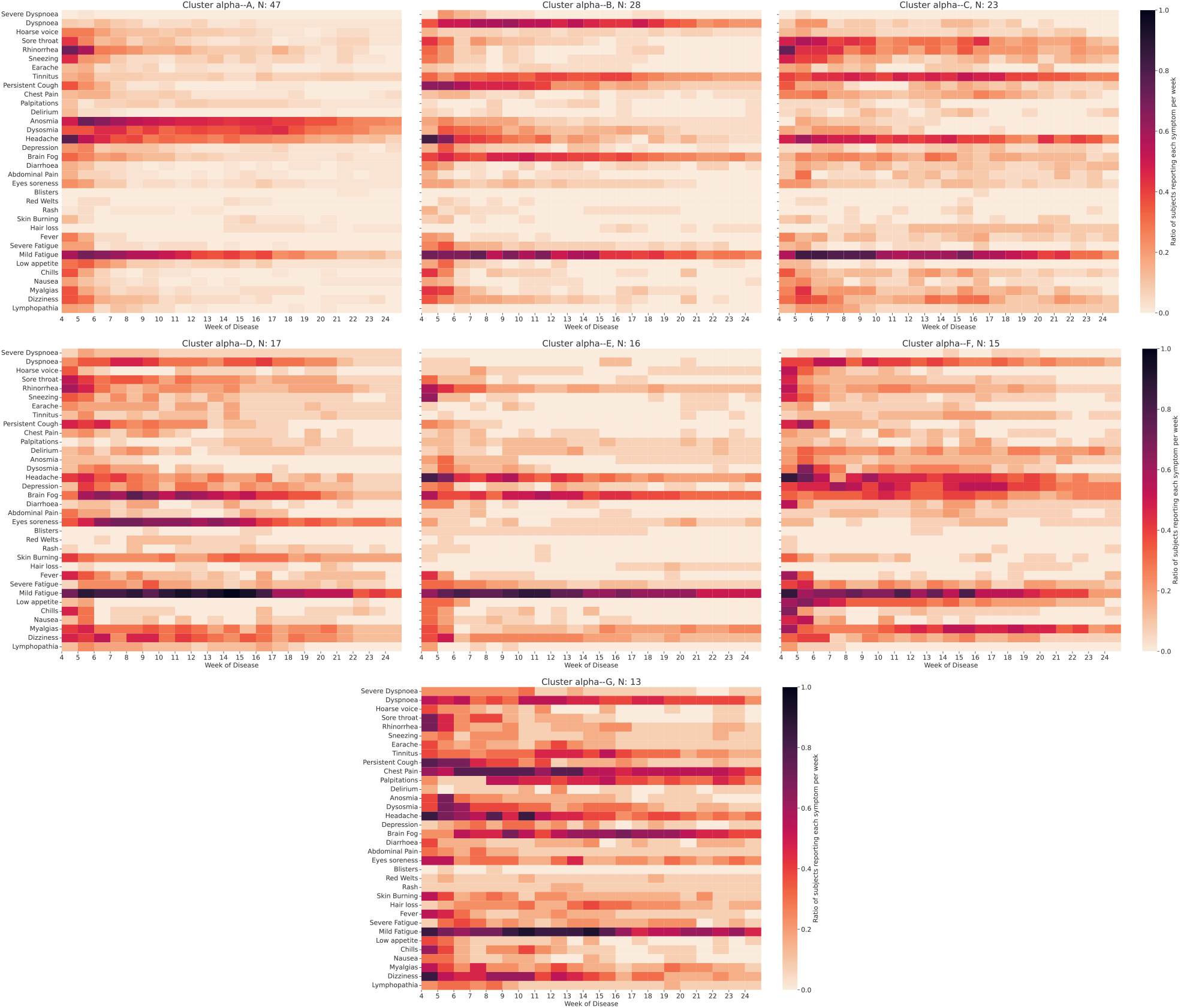
Symptom prevalence profile for PCS clusters for unvaccinated subjects infected with the alpha variant. The proportion of subjects reporting each symptom (ratio) per week is encoded by the colourmap (darker represents a higher ratio).

**Figure 6.**
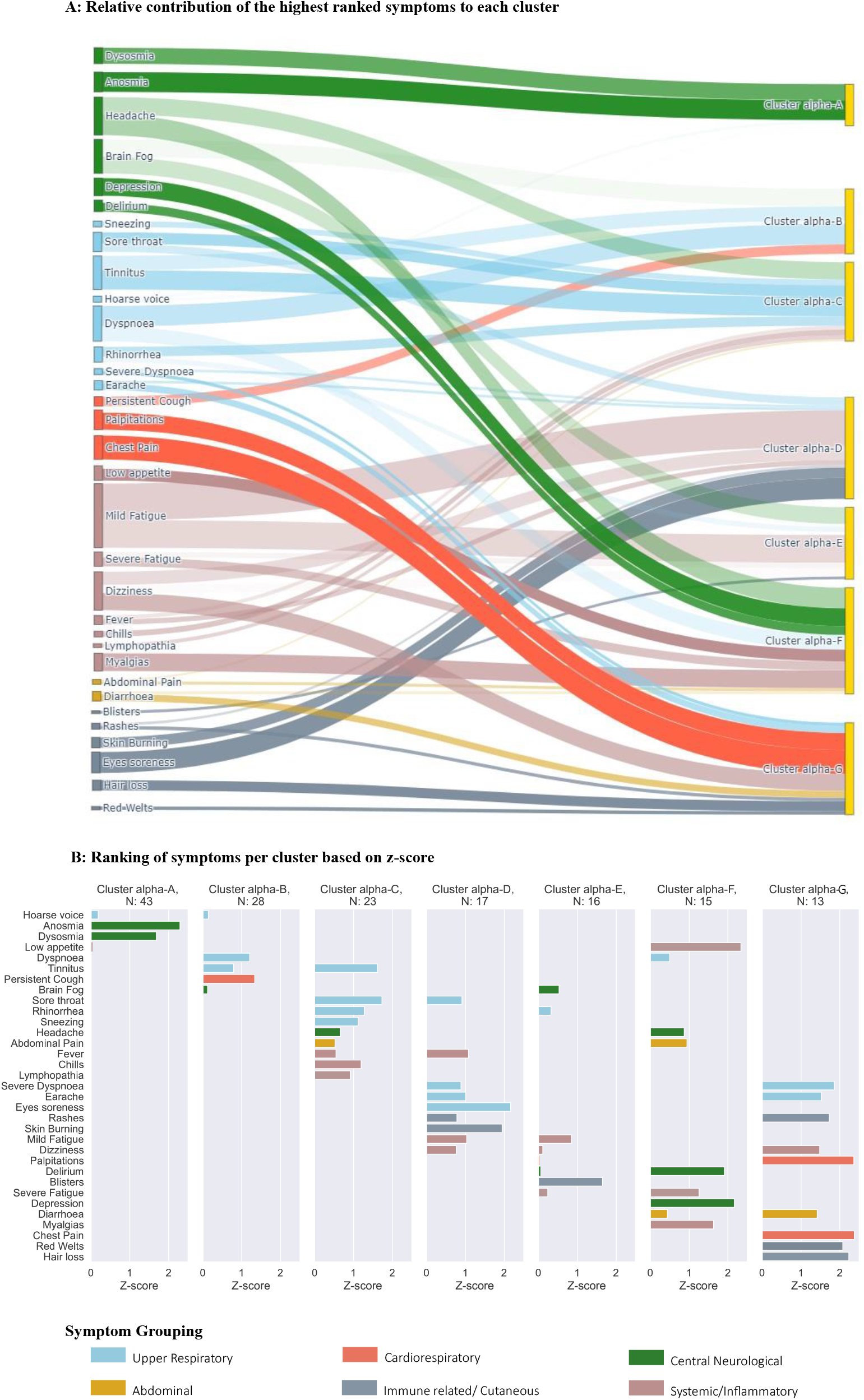
Relative contribution of symptoms per cluster for PCS in unvaccinated individuals infected with the alpha variant. Symptoms are selected based on the 3^rd^ quartile of the z-scores (only positive z-scores shown), aiming to highlight the most influential symptoms in each cluster. **A:** relative contributions of symptoms per cluster. The width of shadow connections encodes the symptom prevalence (median number of individuals reporting the symptom within the cluster), while colour intensity encodes the relevance of symptoms (z-score) per cluster. **B:** Ranking of symptoms based on z-scores, per cluster.

#### PCS Clusters from infection with the delta variant

We identified five clusters of symptoms for vaccinated subjects infected by the delta variant (Figure 7). The largest cluster, cluster delta-A (n=431, 49%) included mainly central neurological symptoms, similar to alpha (Figure 8, Supplementary Figure 9); the smallest cluster, cluster delta-E (80 subjects, 9%), contained predominantly abdominal symptoms. Symptom duration was similar across all clusters (approximately 18-20 weeks after onset – Supplementary Table 4), which was longest with cluster delta-E (median 20 weeks (IQR [16.8; 28.3]), and shortest with cluster delta-C (median 18.0 weeks [14.0; 23.0]). Significant differences were found in BMI between genders for cluster delta-A (Supplementary Figure 11), with a p-value < 0.002, after correction for multiple comparisons. No further demographic differences were found across clusters.

**Figure 7.**
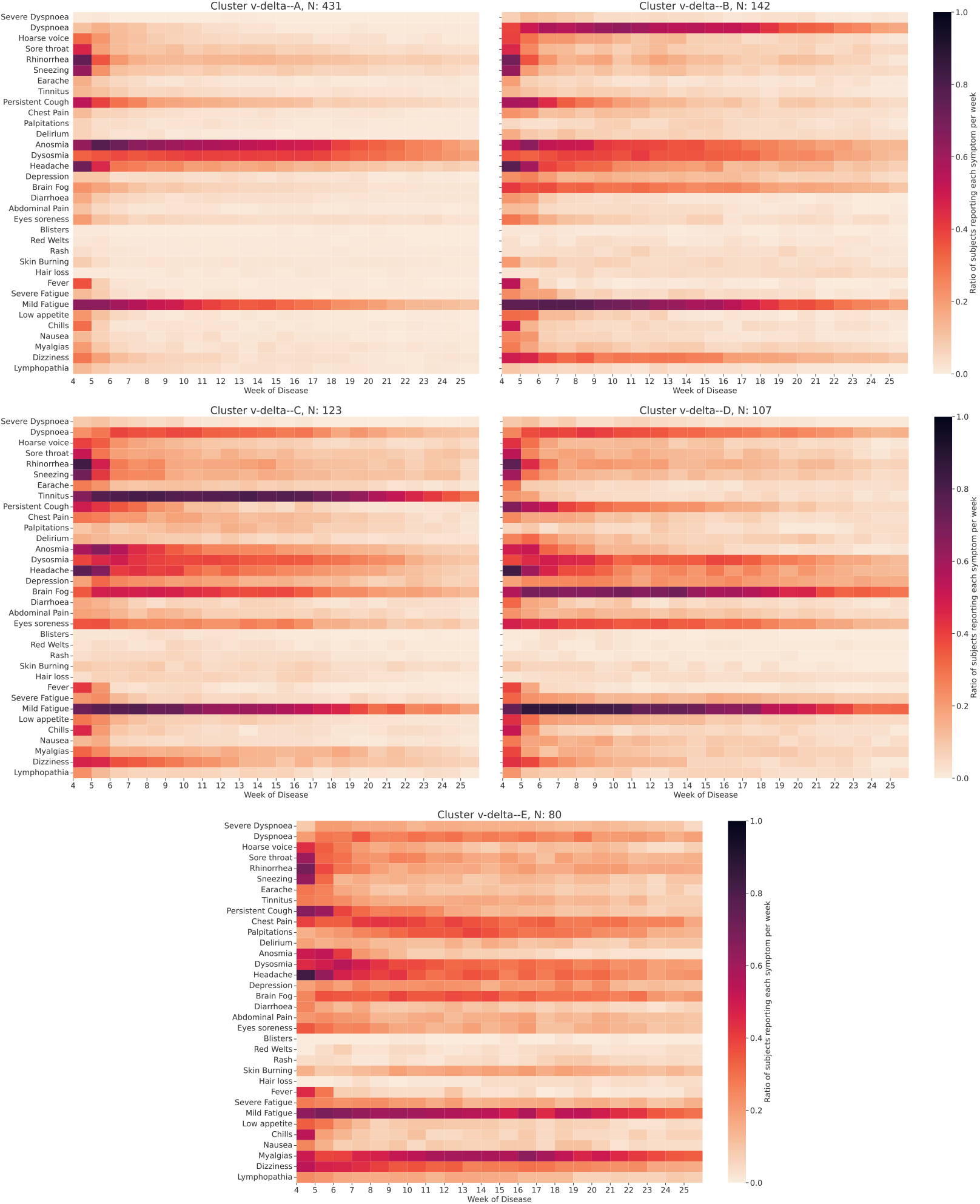
Symptom prevalence profile for PCS clusters for vaccinated individuals infected with the delta variant. The proportion of subjects reporting each symptom (ratio) per week is encoded by the colourmap (darker represents a higher ratio).

**Figure 8.**
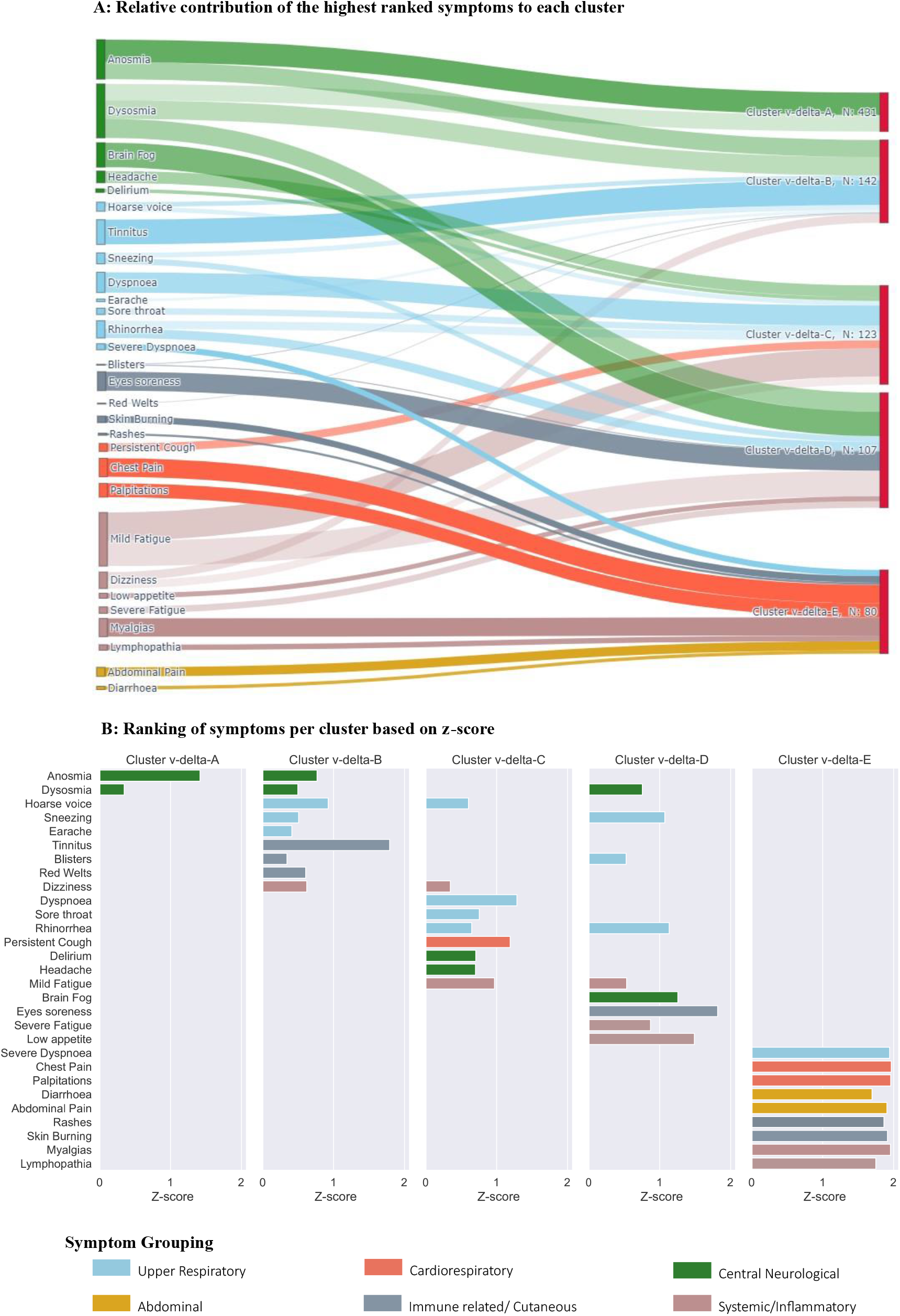
Relative contribution of symptoms per PCS cluster for vaccinated subjects infected with the delta variant. Symptoms are selected based on the 3^rd^ quartile of the z-scores (shown are only positive z-scores), highlighting the most influential symptoms in each cluster. **A:** relative contributions of symptoms per cluster. The width of shadow connections encodes the symptom prevalence (median number of individuals reporting the symptom within the cluster), while colour intensity encodes the relevance of symptoms (z-score) per cluster. **B:** Ranking of symptoms based on z-scores, per cluster.

### Long-lasting effects and clinical validation

We assessed: 1) robustness of the model in predicting the correct cluster for new individuals and (2) relevance of the clusters on clinical outcome of individuals reporting PCS,

Using an independent testing sample, we predicted classification of individuals infected by wild-type SARS-CoV-2. Figure 9 shows symptom prevalence for individuals in the testing sample. Overall, the model correctly classified individuals based on their main symptom profile, according to the clusters found for the wild-type variant (Supplementary Figure 13). Particularly, individuals labelled with cluster wild-test-B showed a similar symptom profile to the population in the training set (high prevalence of mild fatigue, headache, dizziness, dyspnoea and myalgias). We obtained similar results for clusters wild-test-C and wild-test-D, where symptom profiles followed the average profile per cluster derived from the training population. In contrast, the testing cluster wild—test-A appeared qualitatively different from the training cluster wild-A but the error was higher (median error intra-cluster for clusters wild-test-A: 645.99, wild-test-B:448.89, wild-test-C:389.74, wild-test-D:401.03, respectively) and it had the smallest sample size (n = 7), precluding further conclusions.

**Figure 9.**
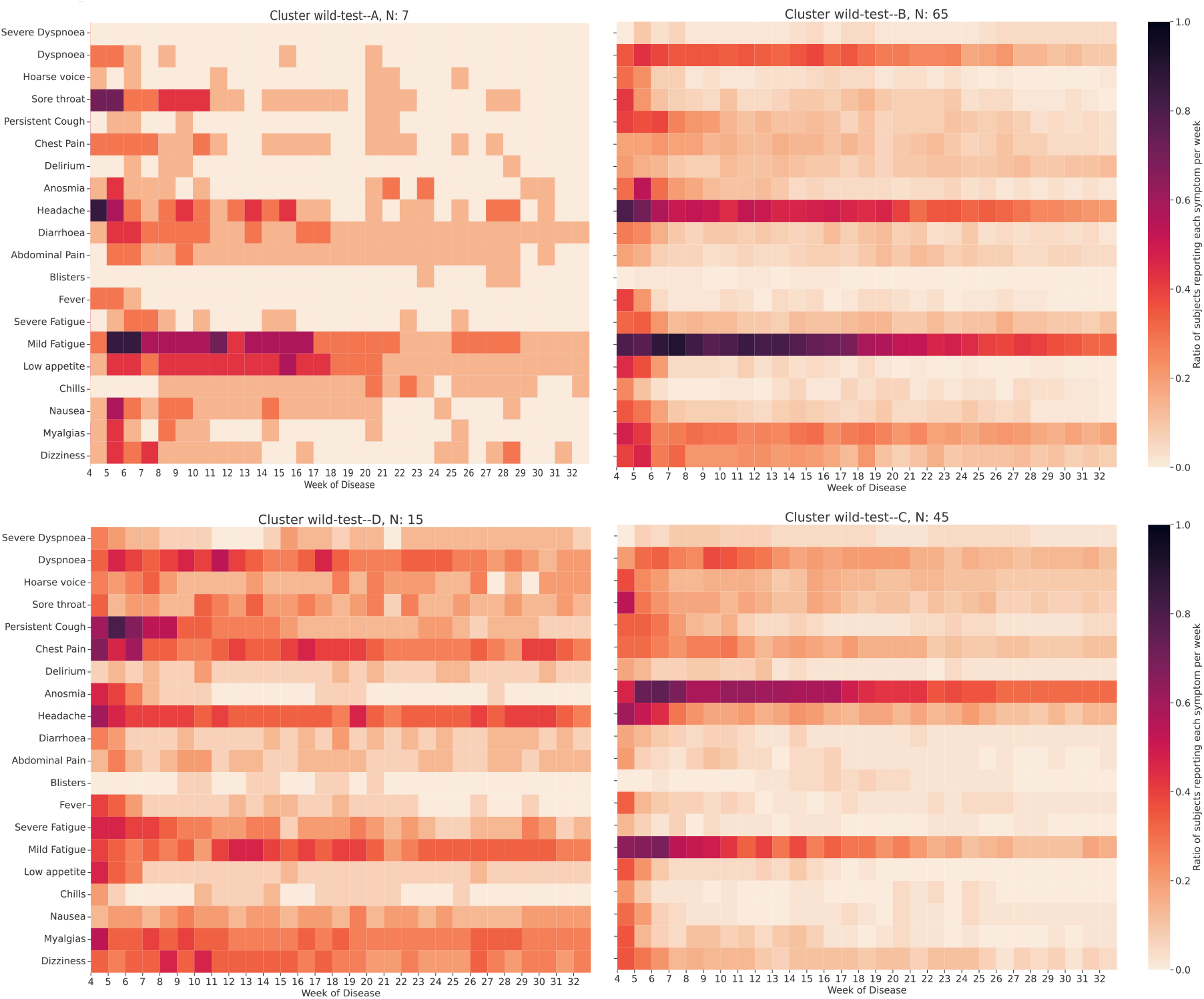
Symptom prevalence profile for the cluster prediction for the independent sample composed unvaccinated individuals infected by the wild-type variant with symptomatic illness for longer than 12 weeks. The proportion of subjects reporting each symptom (ratio) per week is encoded by the colourmap (darker represents a higher ratio).

For the CSSB sample, we assessed the relation between cluster classification and retrospective self-assessments of PCS. Figure 10 shows the odds ratio obtained by a multivariate GLM model (logistic regression), with cluster classification used as a covariate in the model testing associations with healthcare use, re-infection, and impact.

**Figure 10.**
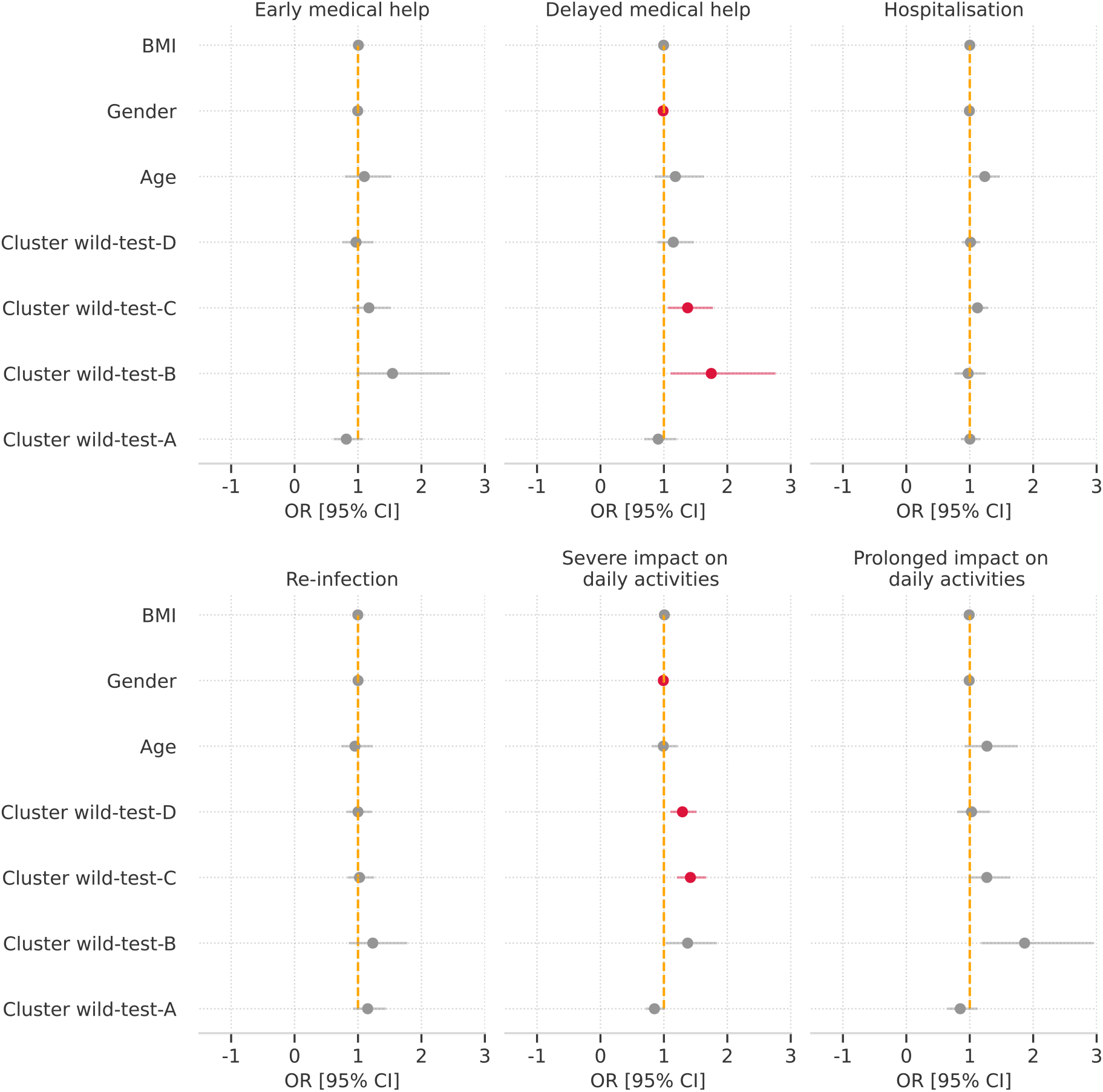
Relation between predicted cluster and live impact of PCS for wild-type variant. Odds ratios (dots) and 95% confidence interval (CI) (error bars) for each variable of the multivariate model are shown. Red encodes significance after false discovery rate (FDR) correction, for alpha = 0.05. Questions reflect the need for acute (< 4weeks), or delayed (>4 weeks) medical help after infection, hospitalisation, reinfection, severe impact on daily life and prolonged impact on daily activities, as summarised in Supplementary Table 2.

Individuals labelled with cluster wild-B had increased odds for illness severity, for both acute and long-lasting symptoms (OR=1.54 [0.91; 2.45], OR=1.75 [1.06; 2.76], for acute and long respectively). Cluster wild-test-C, showed no detectable relation significant (OR= 1.17 [0.75; 1.52], OR=1.37 [0.89; 1.77]). Both clusters wild-B and wild-C contributed significantly to the long severity of symptoms for patients reporting PCS, even after FDR correction. These findings suggest that the presence of respiratory symptoms (severe dyspnoea, hoarse voice), immune-related symptoms (fever), and neurological symptoms (fatigue, delirium, anosmia, and headache) can lead to prolonged severe symptoms in individuals with PCS.

Individuals in clusters wild-test-C and wild-test-D had an increased odds of severe impact on daily life (OR= 1.42 [1.10; 1.67] and OR = 1.29 [0.81; 1.51], respectively), noting that both these clusters are strongly influenced by abdominal and central neurological symptoms, particularly during the first weeks of illness.

Overall, cluster wild-B had the most impactful effect on lives, with an increased likelihood for five out of the six outcomes modelled.

## Discussion

This study profiled phenotypes of PCS for different variants of SARS-CoV-2, using a longitudinal sample of self-reported symptoms. We identified distinct symptom profiles (or clusters) which varied in number and precise symptom combinations between variants, suggesting the existence of heterogeneous profiles of PCS caused by the different SARS-CoV-2 strains. Nevertheless, across variants, there were three groups of symptoms that clustered consistently and reproduced in a hold-out dataset with additional outcome data. Outcome data also demonstrated the association between symptom profile of the optimised clusters and specific daily life difficulties reported by individuals with PCS. These three main clusters of PCS were detectable in all variants of SARS-CoV-2, and included a primary cluster dominated by central neurological symptoms, a second cluster dominated by cardiorespiratory symptoms, and a third more heterogeneous cluster showing systemic and inflammatory symptoms.

Our study is among the first large-scale studies to define in an unsupervised fashion symptom cluster in PCS in adults. Notable previous manuscripts include the Post-hospitalisation COVID-19 (PHOSP) study, which evaluated long-term symptoms in hospitalised patients. This study, like ours, identified several (four) symptom clusters though this was more a gradation of impact: mild physical health impairment; moderate physical health impairment; severe mental and physical health impairment; and very severe mental and physical health impairment. Another study, from the National Core Study for Health and Wellbeing, like our study, incorporated individuals from the general population. This study identified two clusters in individuals more than 12 weeks post infection: high and low symptom burden clusters^37^. This study had many methodological differences to our study. Firstly, all individuals with and without COVID-19 were included in the modelling, of whom few (n=135) reported functional limitation after COVID-19 illness for more than 12 weeks, limiting power to identify different clusters of PCS. Lastly, a different study^38^, focusing the characterization of post-acute sequelae of COVID-19 on hospitalised patients, identified central neurological, cardiorespiratory and systemic/inflammatory, including gastrointestinal disorders, malaise, fatigue, musculoskeletal pain and anaemia.

In the current study, the central neurological cluster was characterised by symptoms such as anosmia/dysosmia, fatigue, brain-fog, depression, delirium and headache. This profile of symptoms corresponded to the largest cluster in both alpha and delta variants, and the second largest for the wild-type variant. Other studies have pointed to neurological involvement in PCS. Notably, UK Biobank (UKBB)^39^ identified significant longitudinal effects when analysing individuals with PCS and comparing their brain imaging data pre and post-infection. The UKBB study demonstrated that individuals developing PCS had a reduction in grey matter thickness, tissue damage in regions functionally connected to the primary olfactory cortex, and a reduction in global brain size. Furthermore, their results focussing on the imaging of the limbic system demonstrated the degenerative spread of the disease via olfactory pathways, which may explain the presence of anosmia/dysosmia as one of the main symptoms of PCS, as identified in our analysis.

The second common cluster was associated with cardiorespiratory symptoms and was the largest cluster in the wild-type period when all individuals were unvaccinated. This cluster may reflect lung damage, identified in other studies and reflected here as dyspnoea and chest pain. Previous studies had also identified cardiorespiratory syndrome with a high prevalence of dyspnoea, fatigue, palpitations, and chest pain^40^. Some studies^41–43^ have demonstrated respiratory damage in some individuals with PCS, explaining these ongoing symptoms^44^.

Lastly, a third common cluster, present in all variants, was distinguished by systemic/inflammatory and abdominal symptoms, and myalgias, as have been reported in previous studies focused on PCS. Other symptoms, reported less frequently distinguished further clusters which differed across variants and vaccination status. While our inability to qualitatively replicate these clusters across variants may indicate our own power limitations, they may also reflect differences in underlying processes related to the variants’ properties. Some of these rarer symptoms have previously been reported in relation to Long COVID phenotypes^40, 45^.

Previous work demonstrated that vaccination reduces severe COVID-19 and hospitalisation^46^ and also the risk of Long COVID ^7, 47^. However, we did not observe evidence of qualitatively different symptom clustering in vaccinated vs. unvaccinated individuals, with either alpha or delta variants. Our data suggested that vaccination status, i.e., pre- or post-vaccination infection, had a marginal effect on the subsequent duration of symptoms, for individuals with PCS. Note that further analyses regarding the time of vaccination, dose and vaccine type were not conducted in this study, preventing additional conclusions regarding vaccination and post-COVID effects.

The clinical relevance of this clustering approach was supported by integration of the clustering results with retrospective questionnaire data about the lived experience of COVID-19. The main result from this analysis demonstrated that the PCS clusters differed in their relation to individuals’ daily activities. Therefore, such clusters could be clinically useful in the future, by providing information to individuals with PCS about their prognosis (including timescale) and likelihood of needing long-term support^37^. Individuals with lived experience of PCS and their families (the CSSB volunteer advisory panel) broadly recognised the characteristics of the clusters presented here, further adding face validity.

### Strengths and Limitations

The primary strength of this work is its uniqueness as the first study aiming to profile PCS, across different SARS-CoV-2 variants, and varying vaccination status. Secondly, the model presented in this study was trained on a large population and subsequently validated with an independent testing sample. Our large sample size also supports the generalizability and robustness of our approach, as do the diverse demographics in our population and the broad spectrum of symptoms, also promote generalisability to other populations.

Our direct discussions with affected individuals were extremely helpful in improving our methodology and enabling us to check whether our findings resonated with real life. Lastly, we also consider that our results may provide meaningful insights to individuals with PCS and enhance their understanding of their illness.

However, we also acknowledge our study’s limitations. CSS app users are not fully representative of the UK population (more middle-aged, more likely to be female, of higher educational status, and over-representative of healthcare workers)^28, 34, 48^. We considered the implications of this concerning the likelihood of an infected person presenting for testing^49^; however, the current work does not address that limitation, given that is focused on PCS and not only on acute COVID-19, which might go under detected when testing is not done. We also acknowledge that different economic and cultural experiences may influence the presentation of COVID-19 and reporting of symptoms^48^. Extending this point, our analyses are UK-based, limiting international generalisability. PCS phenotypes may differ genetically, environmentally and socially and further samples in different populations are needed. We did not conduct any specific analysis considering the ethnicity of the participants as a possible covariate in the model and/or confounding effect. Future work should focus on the validation of the proposed approach in different populations with different demographic features. Comprehensive analyses regarding of vaccination (including timing, doses, and vaccine type) and infection with Omicron variant were not conducted in this study, preventing additional conclusions regarding vaccination and subsequent variant behaviour^5^. Lastly, our model only used the symptoms assessed by direct questioning in the CSS app and we did not consider symptoms reported as free text; thus, we might ignore other symptoms that could modify the clusters.

## Conclusion

We identified distinct self-reported symptom evolutions, or clusters, in individuals with more than 12 weeks of persisting symptoms after SARS CoV-2 infection, using a data-driven approach. Although multiple clusters were found across the three variants assessed, common features emerged for three clusters dominated by certain symptom groups: central neurological, cardiorespiratory, and systemic/inflammatory (Supplementary Table 1). Our findings may have relevance to individuals living with PCS, their health practitioners, and healthcare services, providing data to validate their illness and manage their expectations during its course. This work adds to the emerging evidence that PCS may have sub-types, possibly with differing pathophysiology. For any given individual, their condition may relate to a specific central/neurological process, to respiratory damage, or to a systemic inflammatory cause. Our study suggests that further investigation into mechanisms underlying PCS should consider subdividing affected individuals into different subgroups, which may increase the ability to identify distinct processes underlying these symptom clusters.

## Data Availability

Data collected in the COVID Symptom Study smartphone application are shared with other health researchers through the UK National Health Service-funded Health Data Research UK (HDRUK) and Secure Anonymised Information Linkage consortium, housed in the UK Secure Research Platform (Swansea, UK). Anonymised data are available to be shared with researchers according to their protocols in the public interest

https://web.www.healthdatagateway.org/dataset/fddcb382-3051-4394-8436-b92295f14259

## Acknowledgements

We thank the individuals with PCS who kindly participated in this study, including sharing their opinions regarding this work and how could we improve it to understand the progression and recovery of their symptoms.

ZOE Limited provided in-kind support for all aspects of building, running, and supporting the app and service to all users worldwide. This work is supported by the Wellcome EPSRC Centre for Medical Engineering at King’s College London (WT 203148/Z/16/Z) and the UK Department of Health via the National Institute for Health Research (NIHR) comprehensive Biomedical Research Centre award to Guy’s & St Thomas’ NHS Foundation Trust in partnership with King’s College London and King’s College Hospital NHS Foundation Trust. Investigators also received support from Medical Research Council (MRC) and NIHR (including a grant on Long COVID (COV-LT-0009)), British Heart Foundation (BHF), Alzheimer’s Society, European Union, COVID-19 Driver Relief Fund (CDRF) and the NIHR-funded BioResource, Clinical Research Facility and Biomedical Research Centre (BRC) based at GSTT NHS Foundation Trust in partnership with KCL. SO was supported by the French government, through the 3IA Côte d’Azur Investments in the Future project managed by the National Research Agency (ANR) with the reference number ANR-19-P3IA-0002. This research was funded in part by the Wellcome Trust [215010/Z/18/Z]. For the purpose of Open Access, the author has applied a CC BY public copyright licence to any Author Accepted Manuscript (AAM) version arising from this submission.

## Authors’ contribution

LSC, CS, ELD, MM contributed to study concept and design. LSC, JD contributed to acquisition of data. All the authors had access to the raw data underlying the study. LSC, MM contributed to data analysis and verified the underlying data. LSC, EM, AH, ELD, CS, MM contributed to drafting of the manuscript. All authors contributed to interpretation of data and critical revision of the manuscript. CS and MM contributed to study supervision.

## Data sharing

Data collected in the COVID Symptom Study smartphone application are shared with other health researchers through the UK National Health Service-funded Health Data Research UK (HDRUK) and Secure Anonymised Information Linkage consortium, housed in the UK Secure Research Platform (Swansea, UK). Anonymised data are available to be shared with researchers according to their protocols in the public interest (https://web.www.healthdatagateway.org/dataset/fddcb382-3051-4394-8436-b92295f14259).

## References

1. Worldometer. COVID-19 Coronavirus Pandemic. https://www.worldometers.info/coronavirus/ (accessed June 10, 2020).

2. World Health Organisation. WHO Coronavirus (COVID-19) Dashboard. 2022; published online May 25.

3. NHS UK. Coronavirus (COVID-19). 2020; published online Nov 1. https://www.nhs.uk/conditions/coronavirus-covid-19/ (accessed Feb 2, 2021).

4. Menni C, Valdes AM, Polidori L, et al. Symptom prevalence, duration, and risk of hospital admission in individuals infected with SARS-CoV-2 during periods of omicron and delta variant dominance: a prospective observational study from the ZOE COVID Study. The Lancet 2022; 399: 1618–24.

5. Antonelli M, Pujol JC, Spector TD, Ourselin S, Steves CJ. Risk of long COVID associated with delta versus omicron variants of SARS-CoV-2. The Lancet 2022; 399: 2263–4.

6. NICE: National Institute for Health and Care Excellence. COVID-19 rapid guideline: managing the long-term effects of COVID-19. https://www.nice.org.uk/guidance/NG188. 2021.

7. Office for National Statistics. Prevalence of ongoing symptoms following coronavirus (COVID-19) infection in the UK: 22 June 2022. https://www.ons.gov.uk/peoplepopulationandcommunity/healthandsocialcare/conditionsanddiseases/bulletins/prevalenceofongoingsymptomsfollowingcoronaviruscovid19infectionintheuk/6may2022.2022.

8. Ziauddeen N, Gurdasani D, O’Hara ME, et al. Characteristics and impact of Long Covid: Findings from an online survey. PLOS ONE 2022; 17: e0264331.

9. Sivan M, Parkin A, Makower S, Greenwood DC. Post-COVID syndrome symptoms, functional disability, and clinical severity phenotypes in hospitalized and nonhospitalized individuals: A cross-sectional evaluation from a community COVID rehabilitation service. Journal of Medical Virology 2022; 94: 1419–27.

10. Ayoubkhani D, Pawelek P. Prevalence of ongoing symptoms following coronavirus (COVID-19) infection in the UK 3 February 2022. *medRxiv* 2022.

11. Estiri H, Strasser ZH, Brat GA, et al. Evolving phenotypes of non-hospitalized patients that indicate long COVID. BMC Medicine 2021; 19. DOI:10.1186/s12916-021-02115-0.

12. Yong SJ, Liu S. Proposed subtypes of post-COVID-19 syndrome (or long-COVID) and their respective potential therapies. Reviews in Medical Virology 2021; n/a: e2315.

13. Nurek M, Rayner C, Freyer A, et al. Recommendations for the recognition, diagnosis, and management of long COVID: a Delphi study. British Journal of General Practice 2021; 71: e815.

14. Gavrilova N, Soprun L, Lukashenko M, et al. New Clinical Phenotype of the Post-Covid Syndrome: Fibromyalgia and Joint Hypermobility Condition. *Pathophysiology* 2022; 29: 24–9.

15. Lam GY, Befus AD, Damant RW, et al. Exertional intolerance and dyspnea with preserved lung function: an emerging long COVID phenotype? Respiratory Research 2021; 22: 222.

16. Bierle DM, Aakre CA, Grach SL, et al. Central Sensitization Phenotypes in Post Acute Sequelae of SARS-CoV-2 Infection (PASC): Defining the Post COVID Syndrome. Journal of Primary Care & Community Health 2021; 12: 21501327211030824.

17. Whitfield E, Coffey C, Zhang H, et al. Axes of Prognosis: Identifying Subtypes of COVID-19 Outcomes. AMIA Annu Symp Proc 2022; 2021: 1198–207.

18. Sonnweber T, Tymoszuk P, Sahanic S, et al. Investigating phenotypes of pulmonary COVID-recovery: A longitudinal observational prospective multicenter trial. Elife 2022; 11: e72500.

19. Jesuthasan A, Massey F, Manji H, Zandi MS, Wiethoff S. Emerging potential mechanisms and predispositions to the neurological manifestations of COVID-19. Journal of the Neurological Sciences 2021; 428: 117608.

20. Naeije R, Caravita S. Phenotyping long COVID. European Respiratory Journal 2021; : 2101763.

21. Thygesen JH, Tomlinson C, Hollings S, et al. COVID-19 trajectories among 57 million adults in England: a cohort study using electronic health records. The Lancet Digital Health 2022; 4: e542–57.

22. Caspersen IH, Magnus P, Trogstad L. Excess risk and clusters of symptoms after COVID-19 in a large Norwegian cohort. European Journal of Epidemiology 2022; 37: 539–48.

23. Meza-Torres B DGOCMNASMJGPGBCMCVMEJMGTDB de LS. Differences in clinical presentation with long covid following community and hospital infection, and associations with all-cause mortality: English sentinel network database study . JMIR Public Health and Surveillance 2022; 37668.

24. Fernández-de-las-Peñas C, Martín-Guerrero JD, Florencio LL, et al. Clustering analysis reveals different profiles associating long-term post-COVID symptoms, COVID-19 symptoms at hospital admission and previous medical co-morbidities in previously hospitalized COVID-19 survivors. Infection 2022. DOI:10.1007/s15010-022-01822-x.

25. Reese J, Blau H, Bergquist T, et al. Generalizable Long COVID Subtypes: Findings from the NIH N3C and RECOVER Programs. medRxiv 2022; : 2022.05.24.22275398.

26. Kenny G, McCann K, O’Brien C, et al. Identification of Distinct Long COVID Clinical Phenotypes Through Cluster Analysis of Self-Reported Symptoms. Open Forum Infectious Diseases 2022; 9. DOI:10.1093/ofid/ofac060.

27. Drew DA, Nguyen LH, Steves CJ, et al. Rapid implementation of mobile technology for real-time epidemiology of COVID-19. Science (1979) 2020; 368: 1362–7.

28. Menni C, Valdes AM, Freidin MB, et al. Real-time tracking of self-reported symptoms to predict potential COVID-19. Nature Medicine 2020; 26: 1037–40.

29. Brooks Jackson J, Dempewolf S, Johnson AL, van Ert H, Maury W. Dynamics of Humoral Immunity Post COVID Vaccination – A Case Report. Clin Rev & Cases 2021; 3: 1–2.

30. Zaqout A, Daghfal J, Alaqad I, et al. The initial impact of a national BNT162b2 mRNA COVID-19 vaccine rollout. International Journal of Infectious Diseases 2021; 108: 116–8.

31. Polack FP, Thomas SJ, Kitchin N, et al. Safety and Efficacy of the BNT162b2 mRNA Covid-19 Vaccine. New England Journal of Medicine 2020; 383: 2603–15.

32. COG-UK. COG-UK/Mutation Explorer. https://sars2.cvr.gla.ac.uk/cog-uk/. 2022.

33. Murray B, Kerfoot E, Chen L, et al. Accessible data curation and analytics for international-scale citizen science datasets. Sci Data 2021; 8: 297.

34. Sudre CH, Lee KA, Lochlainn MN, et al. Symptom clusters in COVID-19: A potential clinical prediction tool from the COVID Symptom Study app. Sci Adv 2021; 7: eabd4177.

35. King’s College London. COVID Symptom Study Biobank. https://cssbiobank.com/. 2022; published online May.

36. Li H. Multivariate time series clustering based on common principal component analysis. Neurocomputing 2019; 349: 239–47.

37. Davis HE, Assaf GS, McCorkell L, et al. Characterizing long COVID in an international cohort: 7 months of symptoms and their impact. EClinicalMedicine 2021; 38: 101019.

38. Al-Aly Z, Xie Y, Bowe B. High-dimensional characterization of post-acute sequelae of COVID-19. Nature 2021; 594: 259–64.

39. Douaud G, Lee S, Alfaro-Almagro F, et al. SARS-CoV-2 is associated with changes in brain structure in UK Biobank. Nature 2022; 604: 697–707.

40. Jamal SM, Landers DB, Hollenberg SM, et al. Prospective Evaluation of Autonomic Dysfunction in Post-Acute Sequela of COVID-19. J Am Coll Cardiol 2022. DOI:https://doi.org/10.1016/j.jacc.2022.03.357.

41. Grist JT, Collier GJ, Walters H, et al. Lung Abnormalities Depicted with Hyperpolarized Xenon MRI in Patients with Long COVID. Radiology 2022; : 220069.

42. Farghaly S, Badedi M, Ibrahim R, et al. Clinical characteristics and outcomes of post-COVID-19 pulmonary fibrosis: A case-control study. Medicine 2022; 101: e28639–e28639.

43. Rizvi ZA, Dalal R, Sadhu S, et al. Golden Syrian hamster as a model to study cardiovascular complications associated with SARS-CoV-2 infection. Elife 2022; 11: e73522.

44. Couzin-Frankel J. Clues to Long Covid. Science (1979). 2022; published online June 16.

45. Subramanian A, Nirantharakumar K, Hughes S, et al. Assessment of 115 symptoms for Long COVID (post-COVID-19 condition) and their risk factors in non-hospitalised individuals: a retrospective matched cohort study in UK primary care. Nature Portfolio 2022. DOI:10.21203/rs.3.rs-1343889/v1.

46. Antonelli M, Penfold RS, Merino J, et al. Risk factors and disease profile of post-vaccination SARS-CoV-2 infection in UK users of the COVID Symptom Study app: a prospective, community-based, nested, case-control study. The Lancet Infectious Diseases 2022; 22: 43–55.

47. Venkatesan P. Do vaccines protect from long COVID? Lancet Respir Med 2022; 10: e30.

48. Canas LS, Sudre CH, Capdevila Pujol J, et al. Early detection of COVID-19 in the UK using self-reported symptoms: a large-scale, prospective, epidemiological surveillance study. The Lancet Digital Health 2021; 3: e587–98.

49. Graham MS, May A, Varsavsky T, et al. Knowledge barriers in the symptomatic-COVID-19 testing programme in the UK: an observational study. *medRxiv* 2021; : 2021.03.16.21253719.

