## Supplementary material for "Profiling post-COVID syndrome across different variants of SARS-CoV-2"

**Supplementary Table 1. List of symptom and comorbidity questions asked in the COVID Symptom Study application during the current study period.**

| Symptom | COVID Symptom Study app question | Date of addition to the app | Symptom group |
|---|---|---|---|
| Fever | Fever (at least 37.8C or 100F) | 2020-03-29 | Systemic/inflammatory |
| Persistent Cough | A persistent cough (coughing a lot for more than an hour or 3 or more coughing episodes in 24 hours) | 2020-03-29 | Cardiorespiratory |
| Fatigue | Unusual fatigue (no; mild fatigue; severe fatigue/ I struggle to get out of bed) | 2020-03-29 | Systemic/inflammatory |
| Dyspnoea | Shortness of breath or trouble breathing (no; yes mild symptoms/ slight shortness of breath during ordinary activity: yes significant symptoms/ breathing is comfortable only at rest; yes, severe symptoms/ breathing is difficult even at rest). | 2020-03-29 | Upper respiratory |
| Anosmia/Ageusia | Loss of smell/taste | 2020-03-29 | Central neurological |
| Hoarse Voice | Unusually hoarse voice | 2020-03-29 | Upper respiratory |
| Chest Pain | Unusual chest pain or tightness in your chest | 2020-03-29 | Cardiorespiratory |
| Abdominal Pain | Unusual abdominal pain or stomach-ache | 2020-03-29 | Abdominal |
| Diarrhoea | Diarrhoea | 2020-03-29 | Abdominal |
| Delirium | Confusion, disorientation or drowsiness | 2020-03-29 | Central neurological |
| Eye Soreness | Do your eyes have any unusual eye soreness or discomfort (e.g. light sensitivity, excessive tears, or pink/red eye)? | 2020-04-29 | Immune related/cutaneous |
| Low appetite (anorexia) | Skipping meals | 2020-04-29 | Systemic/inflammatory |
| Headache | Headache | 2020-03-29 | Central neurological |
| Nausea | Nausea or vomiting | 2020-03-29 | Systemic/inflammatory |
| Dizziness | Dizziness or light-headedness | 2020-04-29 | Systemic/inflammatory |
| Sore Throat | Sore or painful throat | 2020-03-29 | Upper respiratory |
| Myalgias | Unusual strong muscle pains or aches | 2020-04-01 | Systemic/inflammatory |

| | | | |
|---|---|---|---|
| Red Welts | Raised, red, itchy welts on the skin or sudden swelling of the face or lips | 2020-11-03 | Immune related/cutaneous |
| Blisters | Red/purple sores or blisters on your feet, including your toes | 2020-04-29 | Immune related/cutaneous |
| Rashes | Rash on your arms or torso | 2020-11-03 | Immune related/cutaneous |
| Sensitive Skin | Strange, unpleasant sensations in your skin like pins & needles or burning | 2020-11-03 | Immune related/cutaneous |
| Hair Loss | Unusual hair loss | 2020-11-03 | Immune related/cutaneous |
| Low mood (depression) | Feeling down, depressed, or hopeless | 2020-11-03 | Central neurological |
| Brain Fog | Loss of concentration or memory (brain fog) | 2020-11-03 | Central neurological |
| Dysosmia/Dysgeusia | Altered smell/taste (things smell or taste different to usual) | 2020-11-03 | Central neurological |
| Rhinorrhoea | Runny nose | 2020-11-03 | Upper respiratory |
| Sneezing | Sneezing more than usual | 2020-11-03 | Upper respiratory |
| Ear Pain | Earache | 2020-11-03 | Upper respiratory |
| Tinnitus | Ringing in your ears | 2020-11-03 | Upper respiratory |
| Lymphadenopathy | Swollen neck glands | 2020-11-03 | Systemic/inflammatory |
| Palpitations | Unusually fast or irregular heartbeat (palpitations) | 2020-11-03 | Cardiorespiratory |
| **Comorbidities** | | | |
| Diabetes | Do you have diabetes? | 2020-06-05 | N/A |
| Cancer | Are you living with cancer? | 2020-06-05 | N/A |
| Lung Disease | Do you have lung disease or asthma? | 2020-06-05 | N/A |
| Heart Disease | Do you have heart disease? | 2020-06-05 | N/A |
| Kidney Disease | Do you have kidney disease? | 2020-06-05 | N/A |

**Supplementary Table 2. List of questions from CSSB Long-COVID questionnaire included in the study.**

| Outcome | Question |
|---|---|
| Early Medical help | Q.B.5 In the first 4 weeks of illness, did you look for any medical help for any symptoms you think may have been caused by COVID-19? |
| Delayed Medical help | Q.B.6 Did you look for any medical help for any symptoms you had more than 4 weeks after your symptoms began, that you think may have been caused by COVID-19? |
| Hospitalisation | Q.B.7 Have you ever had to stay in hospital because of COVID-19 symptoms? |
| Re-infection | Q.B.8 Do you think you have caught COVID-19 more than once? |
| Severe impact on daily activities | Q.B.13 How many days were you or have you been so unwell that you stayed in bed or on the sofa? |
| Prolonged impact in daily activities | Q.B.16 Thinking of how you felt 12 weeks after your COVID-19 illness began, what did you need help with because of COVID-19? |

**Supplementary Table 3. Symptoms prevalence for vaccinated and unvaccinated populations, per SARS-CoV-2 variant.** Mann-Whitney two-sided test was used to assess the differences between the two groups, per variant of the virus. Alpha = 0.05, before multiple corrections. Bold encode significant differences, after multiple corrections (alpha corrected = 0.001).

| | **Alpha Variant** | | | | **Delta Variant** | | | |
|---|---|---|---|---|---|---|---|---|
| **Symptoms** | **Statistics (vaccinated)** | **p-value** | **Number vaccinated** | **Number unvaccinated** | **Statistics (vaccinated)** | **p-value** | **Number vaccinated** | **Number unvaccinated** |
| Persistent Cough | 40111 | 0.331 | 118 | 720 | 177844 | 0.452 | 3798 | 98 |
| Dizziness | 33993 | 0.060 | 114 | 670 | 105051 | 0.977 | 2924 | 72 |
| Palpitations | 5503.5 | 0.995 | 44 | 250 | 9629.5 | 0.412 | 930 | 23 |
| Dyspnoea | 24284.5 | 0.661 | 85 | 555 | 60218 | 0.174 | 2318 | 58 |
| Chest Pain | 16781 | 0.037 | 82 | 478 | 63531.5 | 0.159 | 1914 | 60 |
| Hoarse voice | 22247.5 | 0.805 | 95 | 476 | 83509.5 | 0.953 | 2726 | 61 |
| Sore throat | 38185.5 | 0.030 | 126 | 690 | 157143 | 0.690 | 3485 | 88 |
| Eyes soreness | 18304 | 0.705 | 76 | 469 | 67597.5 | 0.514 | 2258 | 57 |

| | | | | | | | | |
|---|---|---|---|---|---|---|---|---|
| Rhinorrhoea | 62384.5 | 0.020 | 176 | 798 | 261685 | 0.072 | 4737 | 100 |
| Sneezing | 48494 | 0.916 | 146 | 668 | 146331 | 0.360 | 4224 | 65 |
| Earache | 6696 | 0.905 | 48 | 276 | 18323.5 | 0.416 | 1493 | 27 |
| Tinnitus | 11606.5 | 0.291 | 63 | 340 | 26404.5 | 0.529 | 1764 | 28 |
| Fatigue | 103894.5 | 0.814 | 197 | 1066 | 354136 | 0.057 | 5076 | 127 |
| Delirium | 4378.5 | 0.240 | 39 | 254 | 18204.5 | 0.951 | 1145 | 32 |
| Dysomnia | 39057.5 | 0.091 | 116 | 746 | 179083 | 0.964 | 3741 | 96 |
| Anosmia | 24461 | 0.083 | 80 | 546 | 225144 | 0.718 | 4124 | 107 |
| Headache | 86060 | 0.487 | 179 | 994 | 290022.5 | 0.085 | 4730 | 112 |
| Brain Fog | 26892.5 | 0.327 | 100 | 573 | 86311.5 | 0.791 | 2775 | 61 |
| Depression | 16766 | 0.161 | 78 | 477 | 51245 | 0.350 | 1903 | 50 |
| Diarrhoea | 11917 | 0.258 | 61 | 429 | 38476 | 0.310 | 1606 | 44 |
| Abdominal Pain | 11516.5 | 0.931 | 61 | 375 | 28416.5 | 0.623 | 1488 | 40 |
| Low appetite | 18279 | 0.747 | 68 | 525 | 49685 | 0.741 | 1855 | 55 |
| Nausea | 11675 | 0.384 | 58 | 433 | 37538.5 | 0.860 | 1555 | 49 |
| Fever | 20896.5 | 0.054 | 81 | 594 | 84154.5 | 0.150 | 2519 | 74 |
| Chills | 29242 | 0.050 | 108 | 614 | 71373.5 | 0.323 | 2661 | 58 |
| Blisters | 84 | 0.139 | 8 | 32 | 77.5 | 0.095 | 118 | 3 |
| Red Welts | 504.5 | 0.549 | 9 | 100 | 1557.5 | 0.193 | 307 | 8 |
| Lymphopathia | 5496.5 | 0.103 | 46 | 281 | 22497 | 0.175 | 1398 | 37 |
| Rash | 1343.5 | 0.356 | 19 | 125 | 2637 | 0.992 | 351 | 15 |
| Myalgias | 24250 | 0.811 | 82 | 582 | 71218 | 0.348 | 2148 | 62 |
| Skin Burning | 4649.5 | 0.241 | 28 | 293 | 13268 | 0.227 | 866 | 27 |
| Hair loss | 277 | 0.765 | 10 | 59 | 1456.5 | 0.959 | 245 | 12 |

**Supplementary Table 4. Illness description for the post-COVID syndrome across clusters and variants**. Clusters are labelled according to the profile found with the unsupervised algorithm. The number of individuals used to optimise the clusters within a variant (training sample) and the number of individuals classified with each cluster are presented. Median duration, in weeks, and interquartile range [IQR] presented per variant and optimised cluster. Main features and symptoms describe the highest ranked symptoms per cluster, based on z-scores values.

| SARS-CoV-2 variant | Clusters | Number individuals | Duration (weeks) | Main features and symptoms | Classification |
|---|---|---|---|---|---|
| Wild | wild-A | 138 | 24.0 [15.0; 40.8] | ◦ Symptoms groups: cardiorespiratory, central neurological (anosmia) and upper respiratory (dyspnoea).<br>◦ High prevalence of dysosmia.<br>◦ Persistent cough as the most relevant symptom (highest z-score). | Predominantly cardiorespiratory |
| | wild-B | 60 | 27.5 [15.0; 48.3] | ◦ Symptoms groups: systemic/inflammatory, central neurological, upper respiratory, and immune-related/cutaneous.<br>◦ Severe symptoms, namely severe dyspnoea for longer than 8 weeks.<br>◦ High prevalence of systemic symptoms.<br>◦ Unique cluster with immune-related symptoms.<br>◦ Most heterogeneous cluster. | Systemic/inflammatory |
| | wild-C | 38 | 15.0 [13.0; 30.8] | ◦ Symptoms groups: upper respiratory, central neurological and systemic/inflammatory.<br>◦ Hoarse voice as the most relevant symptom (highest z-score).<br>◦ High prevalence of dysosmia, with almost neglectable z-score. | Predominantly respiratory |
| | wild-D | 37 | 22.0 [15.0; 49.0] | ◦ Symptoms groups: upper respiratory, central neurological and abdominal.<br>◦ Diarrhoea and sore throat are the most relevant symptoms (highest z-score).<br>◦ High prevalence of neurological symptoms. | Predominantly central neurological with abdominal |
| | Overall | 273 | 23.0 [15.0; 43.0] | ◦ Three main clusters: (1) with cardiorespiratory symptoms, (2) systemic/inflammatory and (3) central neurological.<br>◦ Abdominal and cutaneous symptoms isolated in singular clusters, with high relevance but lower prevalence. | --- |
| Alpha | alpha-A | 43 | 18.0 [14.0; 35.0] | ◦ Symptoms groups: upper respiratory, central neurological and cardiorespiratory.<br>◦ Anosmia is the most relevant symptom.<br>◦ High prevalence of central neurological symptoms. | Central neurological |

| | | | | | |
|---|---|---|---|---|---|
| | alpha-B | 28 | 16.5 [14.0; 28.8] | ◦ Symptoms groups: upper respiratory, central neurological and cardiorespiratory.<br>◦ Persistent cough is the most relevant symptom, followed by dyspnoea.<br>◦ High prevalence of respiratory symptoms and persistent cough. | Predominantly upper respiratory with cardiorespiratory. |
| | alpha-C | 23 | 22.0 [15.5; 55.0] | ◦ Symptoms groups: upper respiratory, central neurological, systemic/inflammatory and abdominal.<br>◦ Strong relevance of respiratory symptoms, followed by systemic symptoms.<br>◦ High prevalence of respiratory symptoms and persistent cough. | Predominantly upper respiratory with systemic. |
| | alpha-D | 17 | 17.0 [14.0; 43.0] | ◦ Symptoms groups: upper respiratory, systemic/inflammatory and immune-related/cutaneous.<br>◦ Strong relevance of respiratory symptoms, followed by immune-related symptoms.<br>High prevalence of systemic and immune-related symptoms. | Predominantly immune-related/cutaneous with systemic. |
| | alpha-E | 16 | 23.0 [13.7; 41.0] | ◦ Symptoms groups: upper respiratory, systemic/inflammatory, immune-related/cutaneous, cardiorespiratory and central neurological.<br>◦ Strong relevance of immune-related symptoms, namely blisters.<br>◦ High prevalence of mild fatigue.<br>◦ Highly heterogeneous cluster. | Predominantly mild symptoms. |
| | alpha-F | 15 | 18.0 [14.5; 37.5] | ◦ Symptoms groups: upper respiratory, systemic/inflammatory, abdominal and central neurological.<br>◦ Strong relevance of systemic/inflammatory followed by neurological.<br>◦ High prevalence of central neurological and systemic symptoms. | Predominantly central neurological with systemic/inflammatory. |
| | alpha-G | 13 | 21.0 [15.0; 44.0] | ◦ Symptoms groups: upper respiratory, systemic/inflammatory, immune-related, abdominal and cardiorespiratory.<br>◦ Strong relevance of cardiorespiratory, followed by immune-related symptoms.<br>◦ High prevalence of cardiorespiratory symptoms. | Multi-systemic with the predominance of cardiorespiratory symptoms. |
| | Overall | 155 | 18.0 [14.0; 43.5] | ◦ Three main clusters: (1) with cardiorespiratory symptoms, (2) systemic/inflammatory and | --- |

| | | | | | |
|---|---|---|---|---|---|
| | | | | immune-related and (3) central neurological.<br>Abdominal symptoms influence mostly the smaller cluster and immune-related symptoms are highly prevalent in clusters with systemic symptoms. | |
| Delta | v-delta-A | 431 | 18.0 [14.0; 24.0] | ◦ Symptoms groups: central neurological.<br>◦ Strong relevance of anosmia with high prevalence. | Central neurological |
| | v-delta-B | 142 | 19.0 [14.0; 23.8] | ◦ Symptoms groups: upper respiratory, systemic/inflammatory, immune-related, and central neurological.<br>◦ Strong relevance immune-related symptoms, namely blisters.<br>◦ High prevalence of upper respiratory and central neurological symptoms. | Predominantly upper respiratory with central neurological. |
| | v-delta-C | 123 | 18.0 [14.0; 23.0] | ◦ Symptoms groups: upper respiratory, systemic/inflammatory, central neurological and cardiorespiratory.<br>◦ Strong relevance of upper respiratory, followed by cardiorespiratory.<br>◦ High prevalence of mild fatigue. | Multi-systemic with a predominance of mild fatigue, upper respiratory and headache. |
| | v-delta-D | 107 | 20.0 [16.0; 25.0] | ◦ Symptoms groups: upper respiratory, systemic/inflammatory, central neurological and immune-related/cutaneous.<br>◦ Strong relevance of immune-related, followed by systemic/inflammatory.<br>◦ High prevalence of mild fatigue and neurological symptoms. | Predominantly immune-related with central neurological and systemic. |
| | v-delta-E | 80 | 20.0 [16.8; 28.3] | ◦ Symptoms groups: upper respiratory, systemic/inflammatory, immune-related/cutaneous, abdominal and cardiorespiratory.<br>◦ Strong influence of cardiorespiratory symptoms and severe dyspnoea.<br>◦ High prevalence of mild fatigue and neurological symptoms. | Severe symptoms |
| | Overall | 155 | 19.0 [15.0; 24.0] | ◦ Three main clusters: (1) with cardiorespiratory symptoms – severe symptoms, (2) systemic/inflammatory and immune-related and (3) central neurological.<br>◦ Abdominal symptoms influence mostly the smaller cluster and immune-related symptoms are highly prevalent in clusters with systemic symptoms. | --- |

**Supplementary Figure 1. SARS-CoV-2 variants of concern in the UK by weeks, from 15th of March 2020 to 3rd of March 2022.** Figure adapted from COG-UK/Mutation Explorer[32]. The variant periods are defined when the prevalence of the variant is higher than 80%. Variant sub-lineages are neglected and analysed as a single variant.

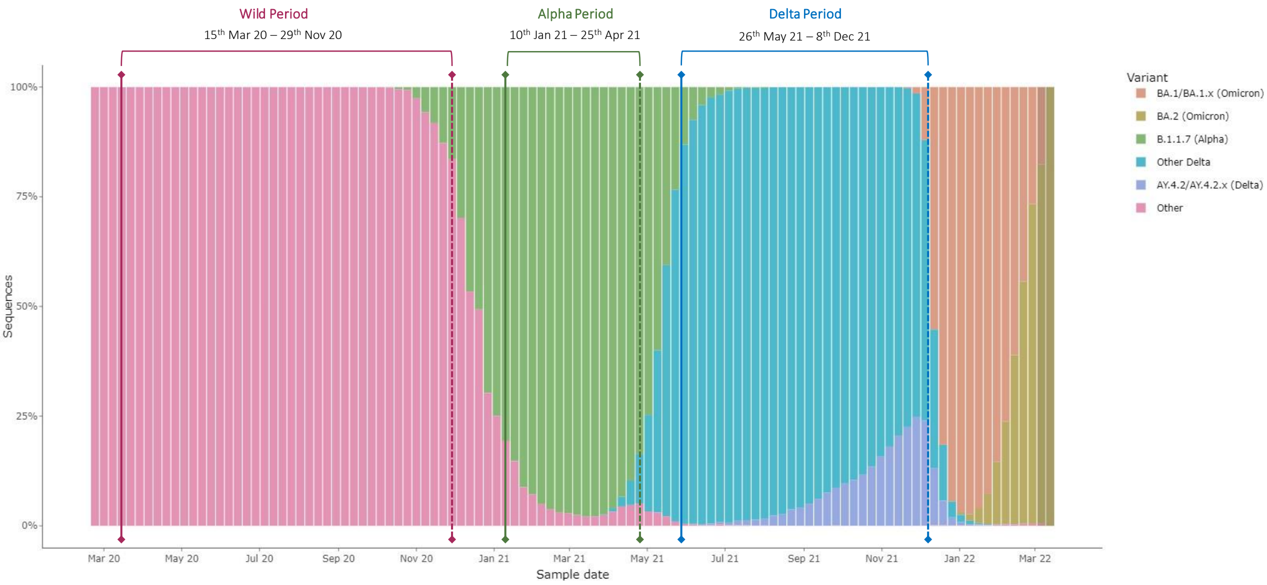


**Supplementary Figure 2. Optimisation of BIC to select the best number of clusters per SARS-CoV-2 variant.** A: Bootstrapping analysis for 50 randomly selected populations for training and testing set, for wild-type variant. B: Bootstrapping analysis for 20 randomly selected populations for training for alpha (left panel) and delta (right panel) variants. Panel B assesses the robustness of the optimised number of clusters. Given early convergence and a smaller sample to bootstrap (small sample size of CSSB sample with both alpha- and delta variants), only 20 runs were performed. No formal testing was performed for these samples.

**A: Wild-type training, 50 runs** **B: alpha-type and delta-type training, 20 runs**

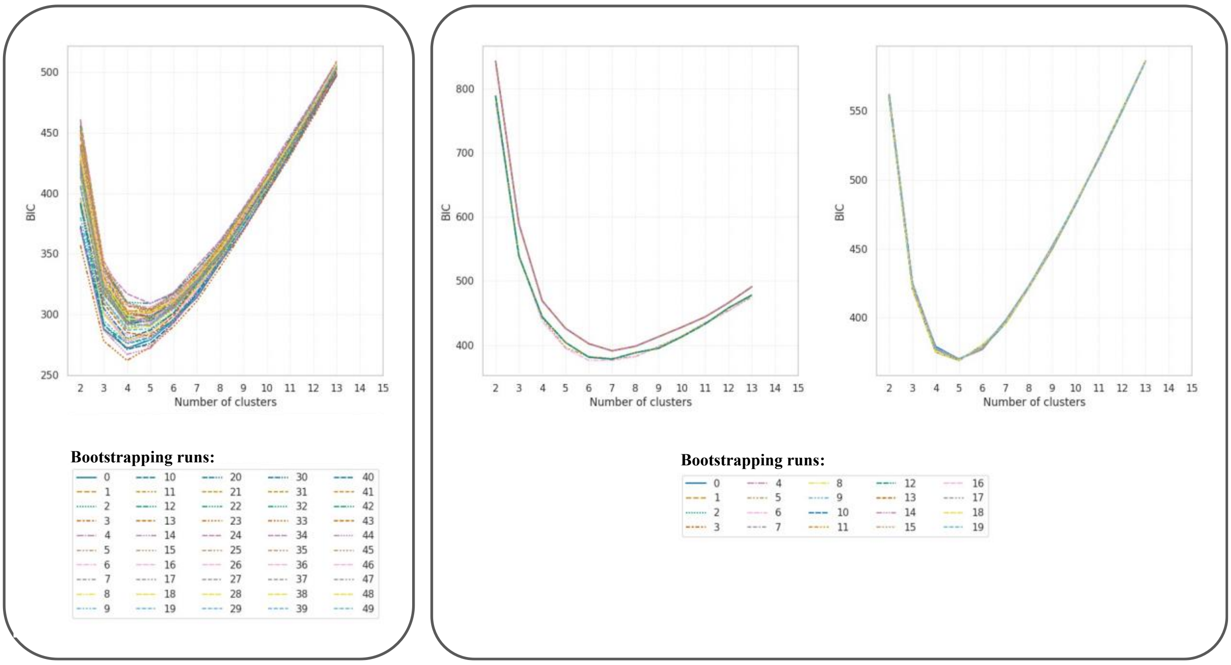

**Supplementary Figure 3. Significance analysis of symptoms profiles per cluster for the wild-type variant of the virus, for symptomatic disease longer than 12 weeks.**

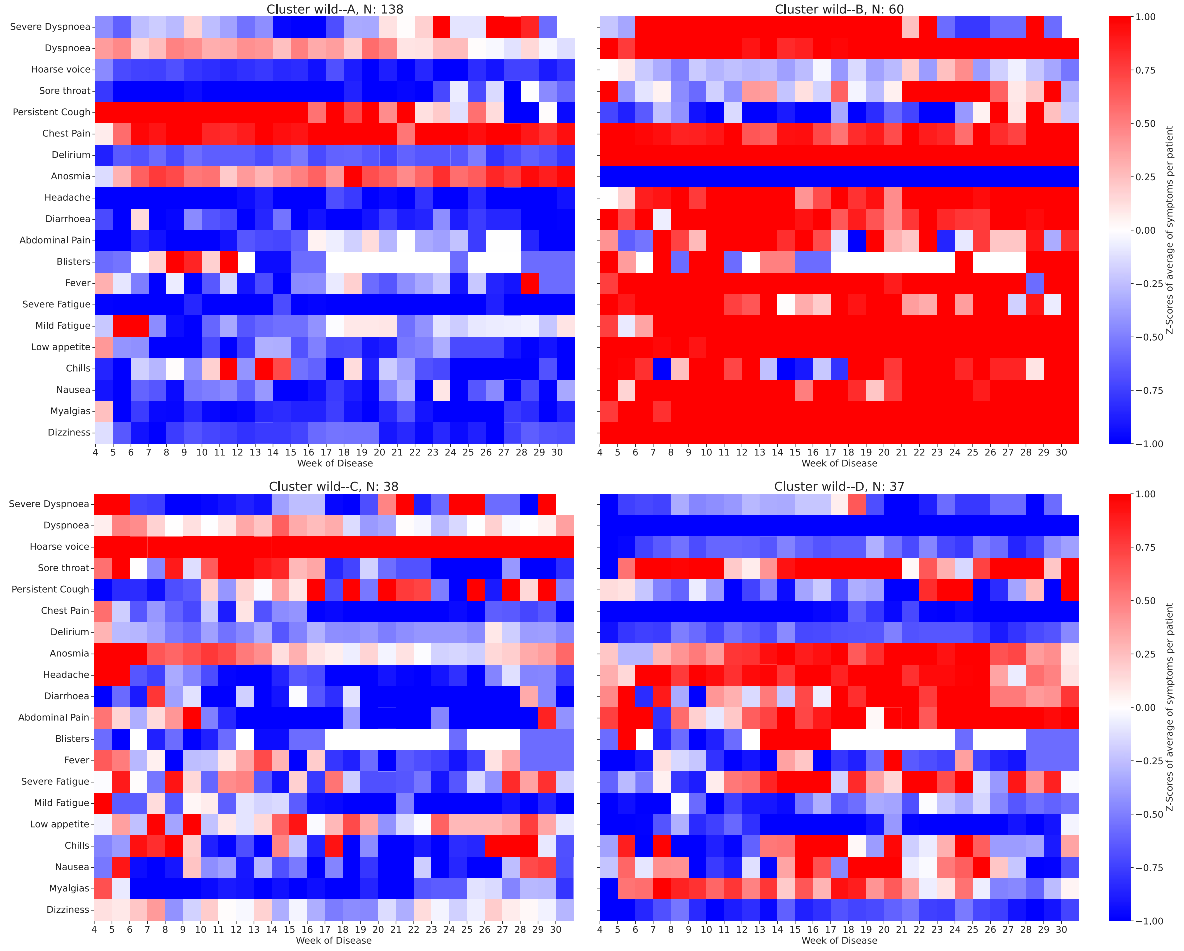


**Supplementary Figure 4. Demographic profile per cluster for the wild-type variant of the virus, for symptomatic disease longer than 12 weeks.** A: Body Mass Index (BMI) in kg/m$^2$; B: Age in years. The diamond (♦) encodes outliers. Mann-Whitney test was conducted to assess statistical differences, alpha = 0.05.

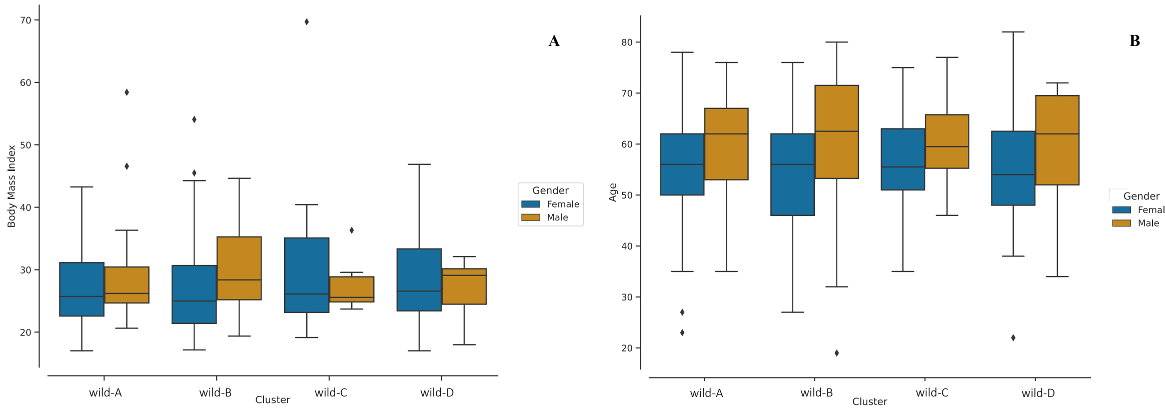

**Supplementary Figure 5. Individual profile for the clusters optimised for the wild-type variant for subjects with the symptomatic disease for longer than 12 weeks.** The duration of each symptom is presented by the colour bar. The grey dots encode the average duration of each symptom for the population labelled with each cluster. The gradient colours encode the prevalence of the symptoms per week – darker encodes a higher prevalence of the symptom.

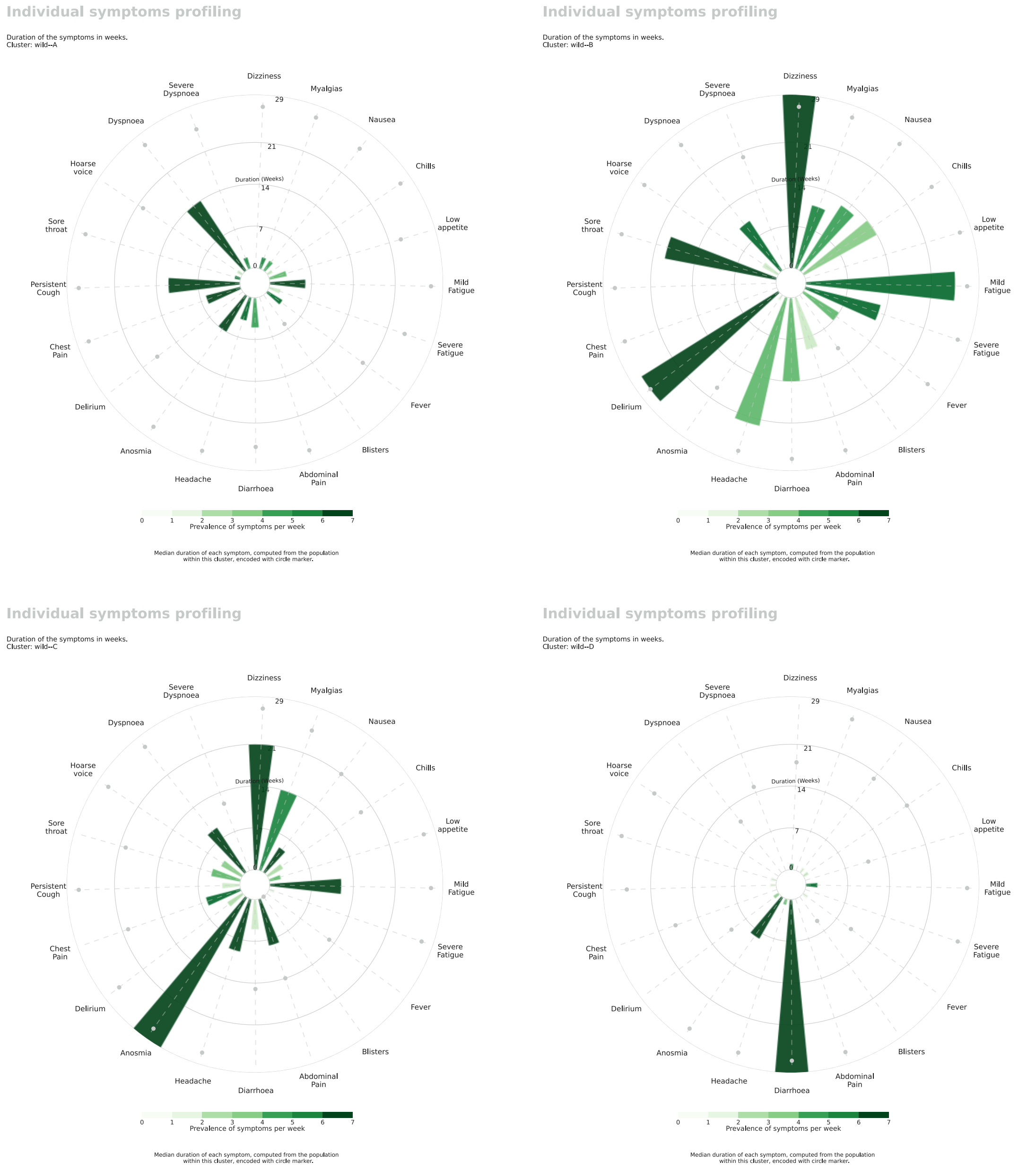

**Supplementary Figure 6. Significance analysis of symptoms profiles per cluster for the alpha variant of the virus, for symptomatic disease longer than 12 weeks.**

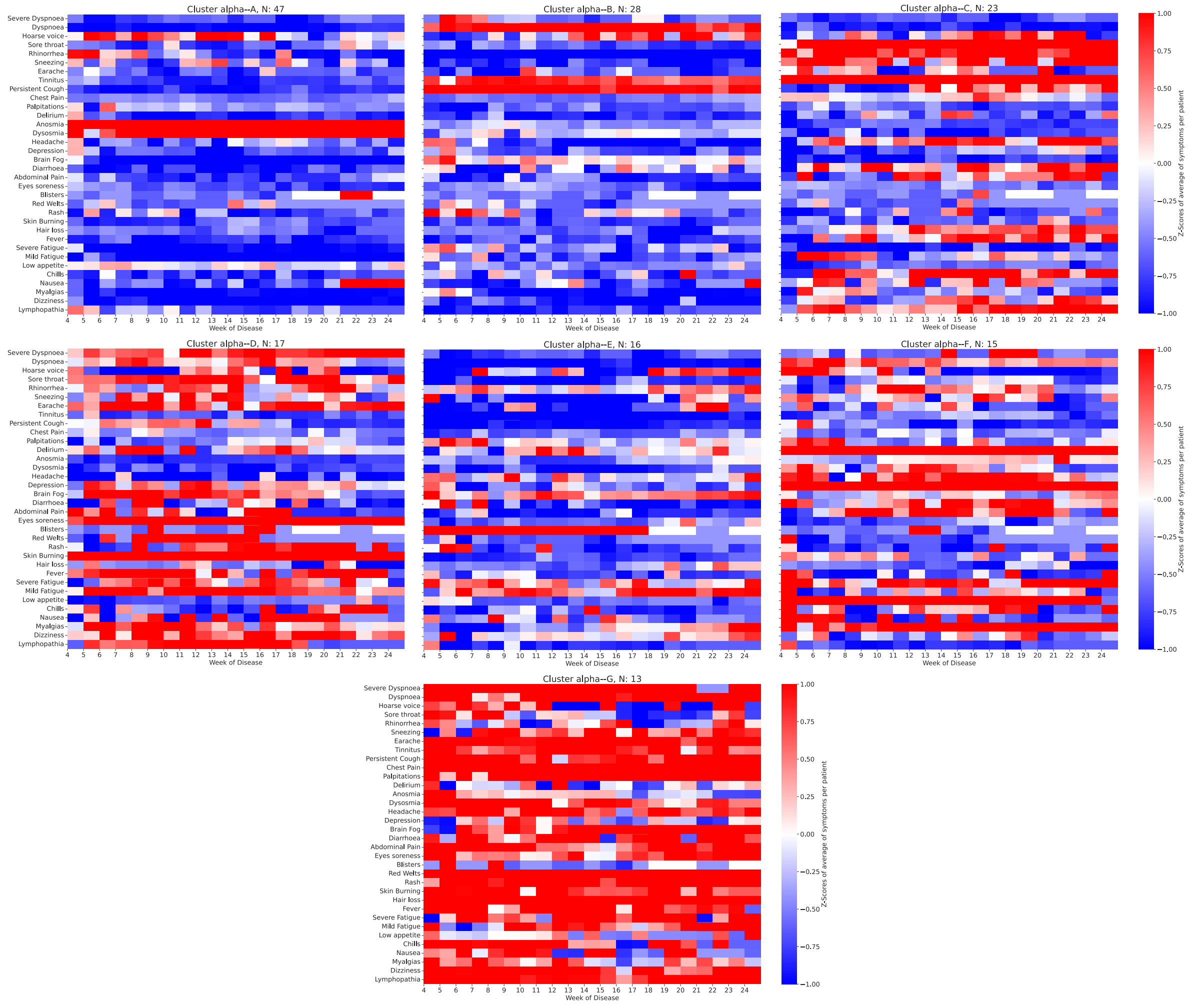

**Supplementary Figure 7. Individual profile for the clusters optimised for the unvaccinated alpha variant for subjects with the symptomatic disease for longer than 12 weeks.** The duration of each symptom is presented by the colour bar. The grey dots encode the average duration of each symptom for the population labelled with each cluster. The gradient colours encode the prevalence of the symptoms per week – darker encodes a higher prevalence of the symptom.

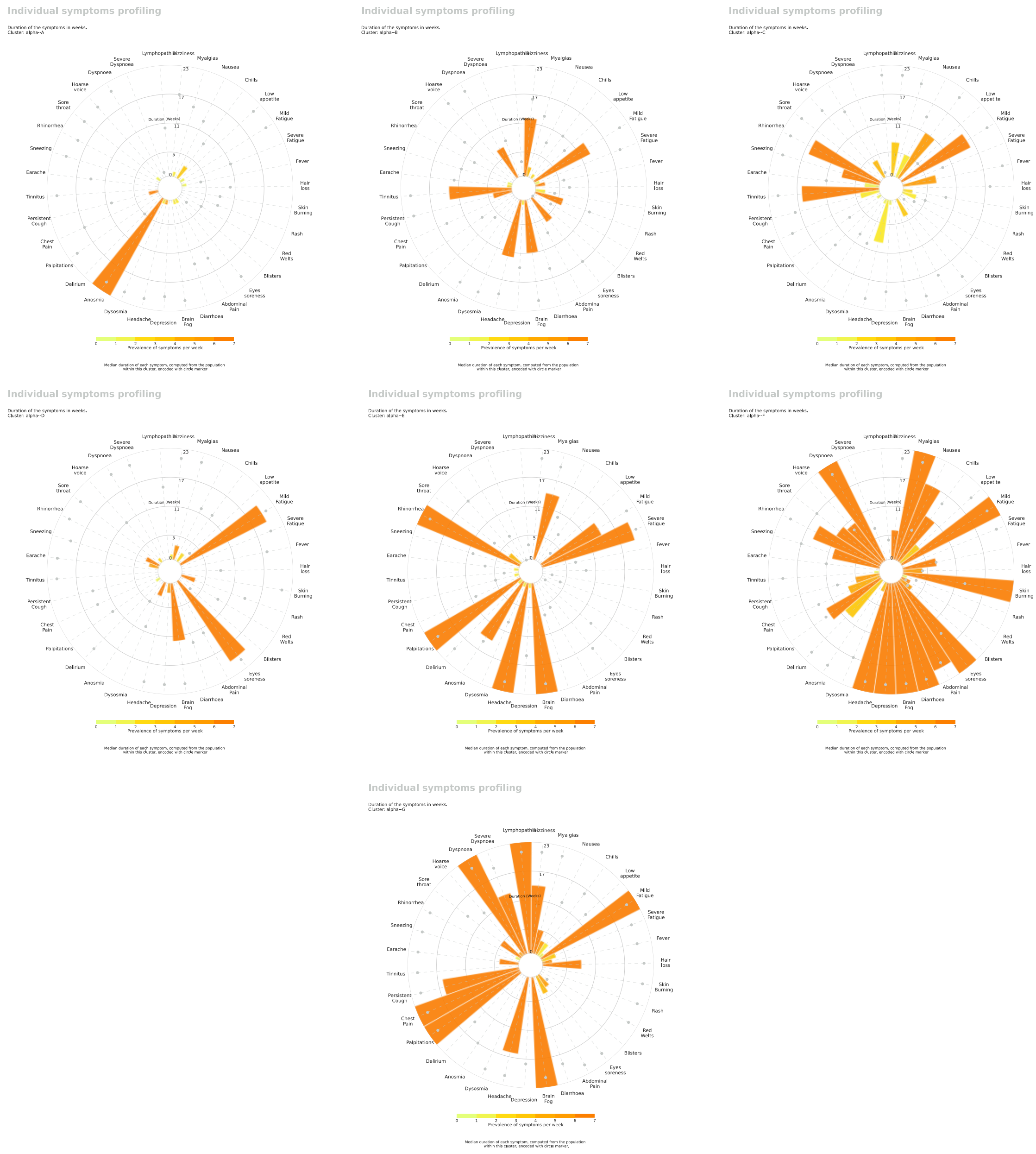

**Supplementary Figure 8. Demographic profile per cluster for the unvaccinated alpha variant of the virus, for symptomatic disease longer than 12 weeks.** A: Body Max Index (BMI) in kg/m$^2$; B: Age in years. The diamond (♦) encodes outliers. Mann-Whitney test was conducted to assess statistical differences, alpha = 0.05.

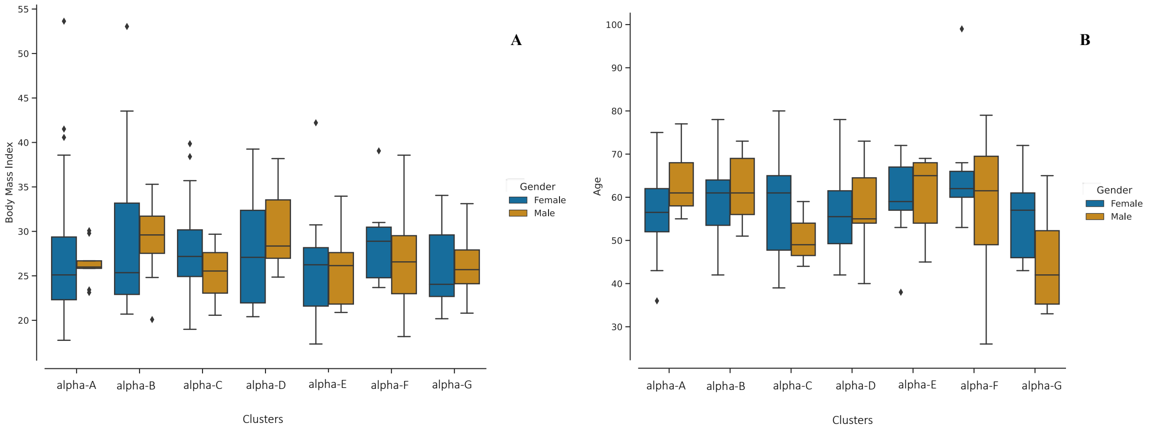

**Supplementary Figure 9. Significance analysis of symptoms profiles per cluster for the vaccinated delta variant of the virus, for symptomatic disease longer than 12 weeks, for vaccinated subjects.**

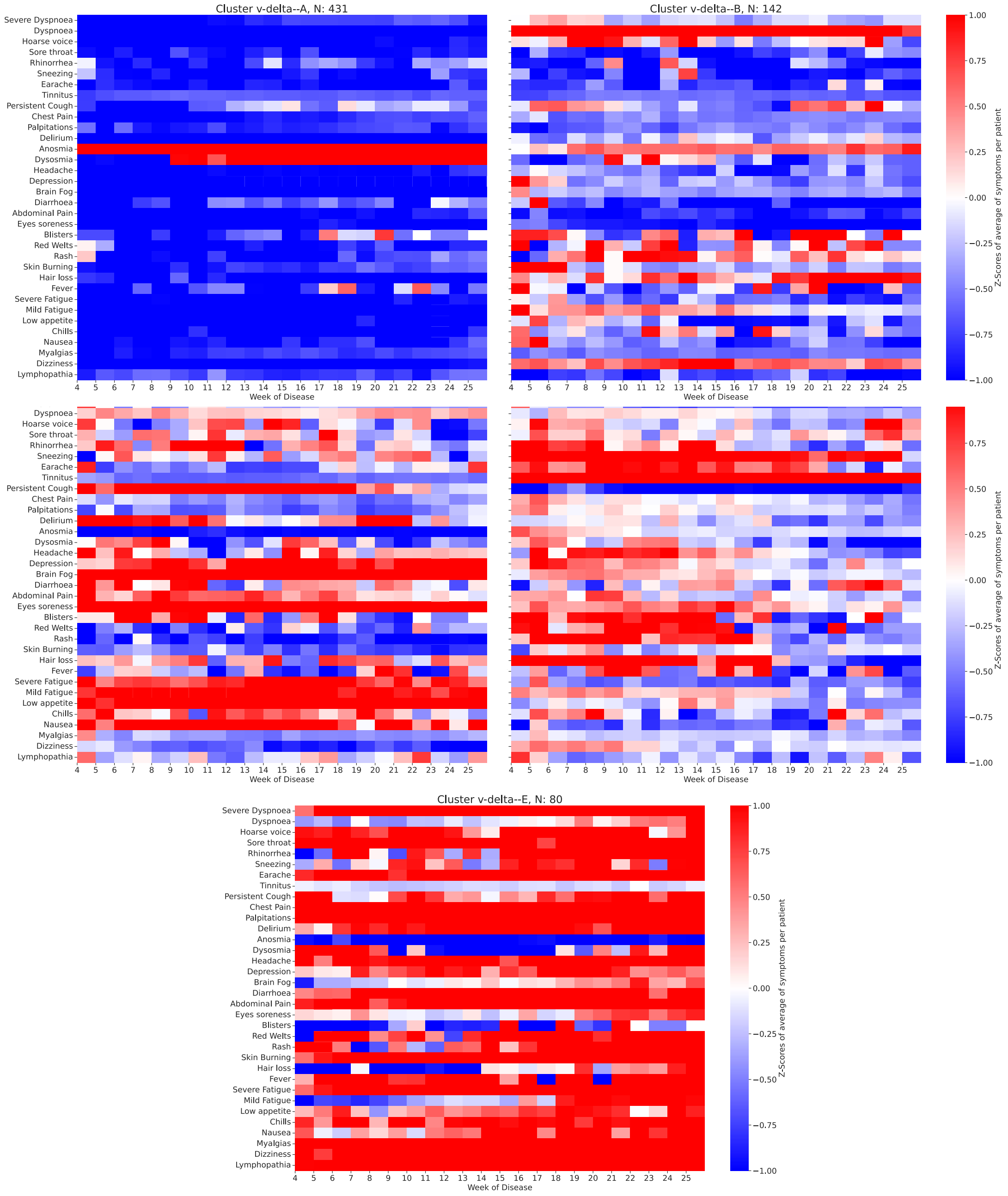

**Supplementary Figure 10. Individual profile for the clusters optimised for vaccinated delta variant for subjects, who were infected after vaccination, with symptomatic lasting for longer than 12 weeks.** The duration of each symptom is presented by the coloured bar. The grey dots encode the average duration of each symptom for the population labelled with each cluster. The gradient colours encode the prevalence of the symptoms per week – darker encodes a higher prevalence of the symptom.

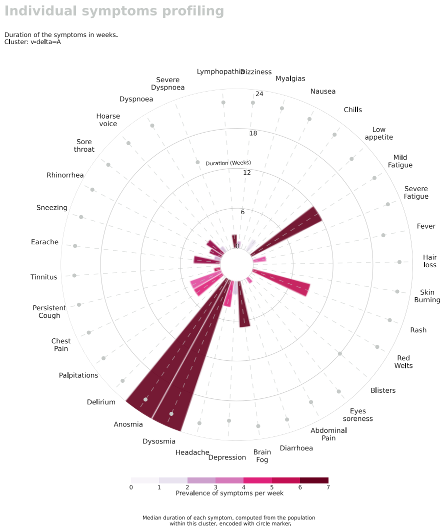


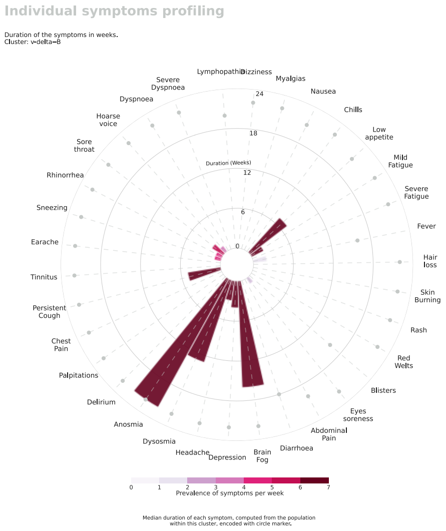


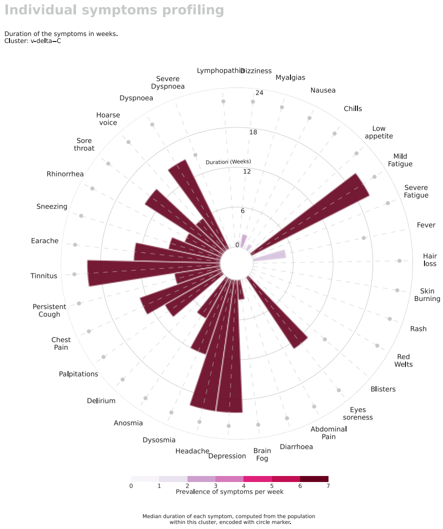


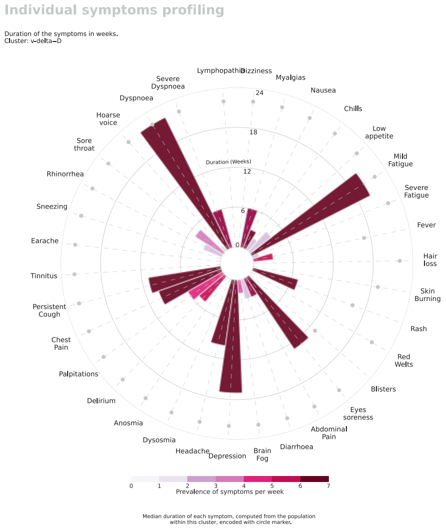


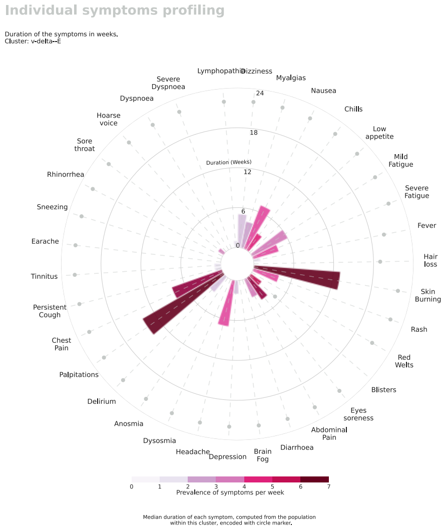

**Supplementary Figure 11. Demographic profile per cluster for the vaccinated delta variant of the virus, for symptomatic disease longer than 12 weeks.** A: Body Max Index (BMI) in kg/m$^2$; B: Age in years. The diamond (♦) encodes outliers. Mann-Whitney test was conducted to assess statistical differences, alpha = 0.05. Asterisk (*) denotes statistical significance.

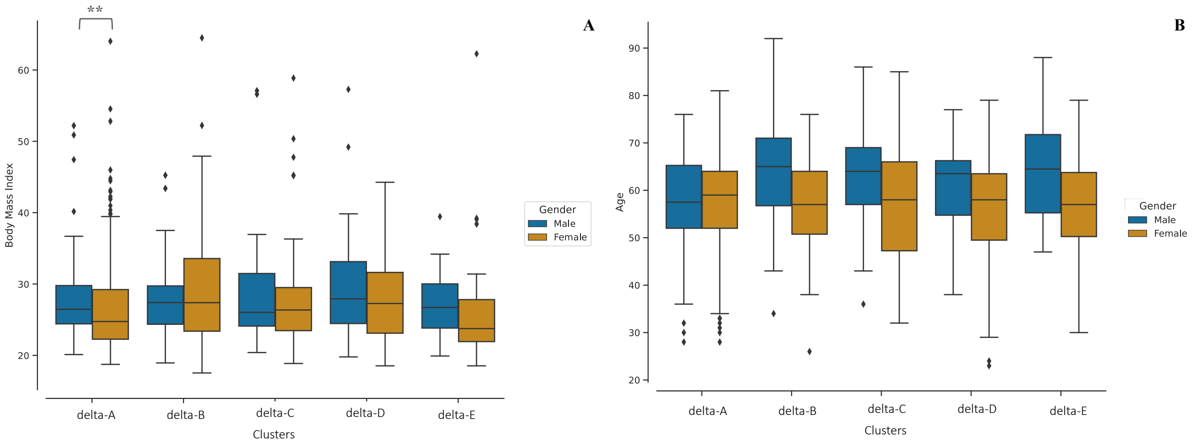

**Supplementary Figure 12. Comorbidity profile per cluster for the three variants of the virus, for symptomatic disease longer than 12 weeks.**

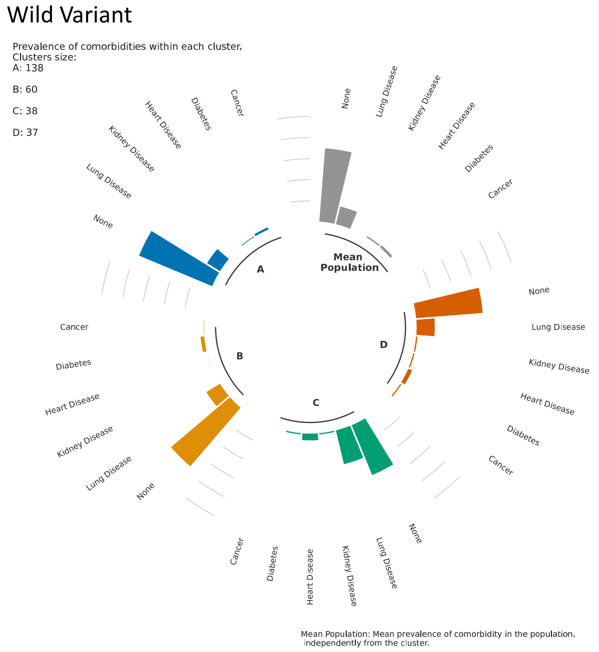


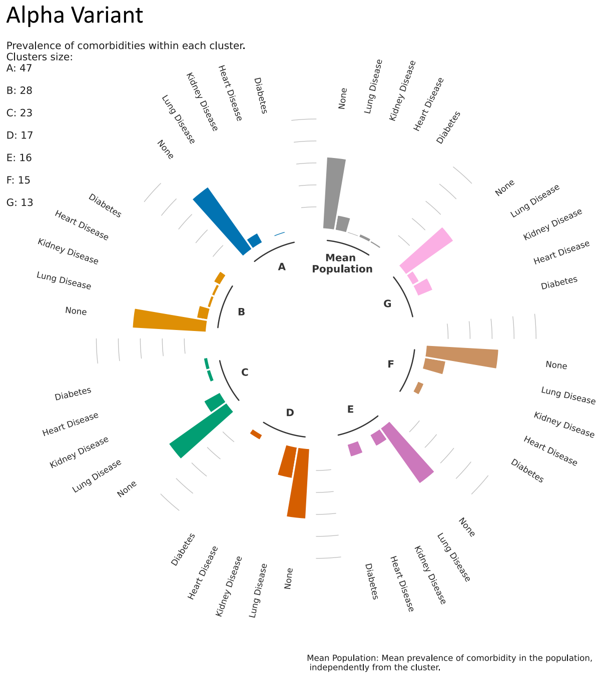


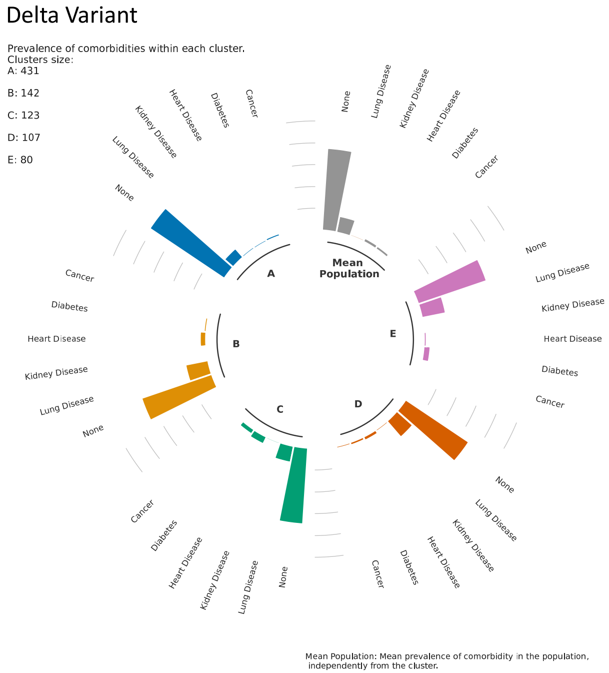

**Supplementary Figure 13. Similarity matrix between the symptom profile of the predicted clusters (wild-type variant) and the symptom profile of training set (wild-type variant).** Median ratio of individuals reporting a symptom across illness is used to identify the top symptoms per cluster in the testing set (right column). The Euclidian distance between the top four (20%) symptom profile per testing cluster and equivalent symptom profile per training cluster is computed as similarity measure. Small differences identify the best matching symptom profile during the illness.

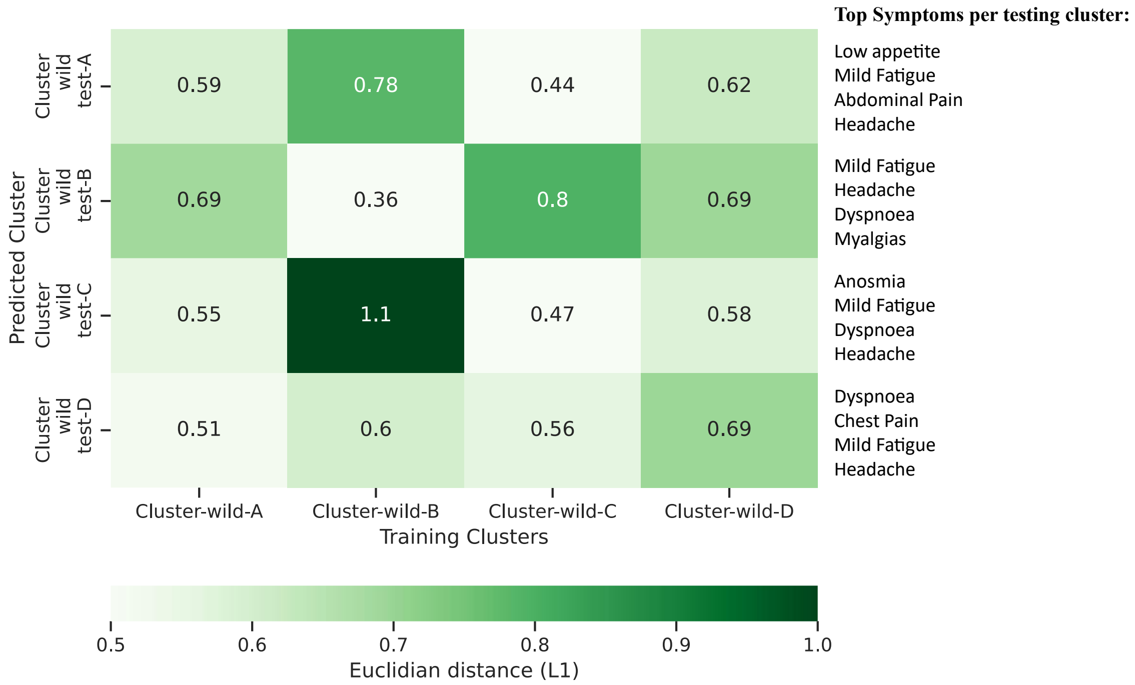
